# Forecasting COVID-19 activity in Australia to support pandemic response: May to October 2020

**DOI:** 10.1101/2022.08.04.22278391

**Authors:** Robert Moss, David J. Price, Nick Golding, Peter Dawson, Jodie McVernon, Rob J. Hyndman, Freya M. Shearer, James M. McCaw

## Abstract

As of January 2021, Australia had effectively controlled local transmission of COVID-19 despite a steady influx of imported cases and several local, but contained, outbreaks in 2020. Throughout 2020, state and territory public health responses were informed by weekly situational reports that included an ensemble forecast for each jurisdiction. We present here an analysis of one forecasting model included in this ensemble across the variety of scenarios experienced by each jurisdiction from May to October 2020. We examine how successfully the forecasts characterised future case incidence, subject to variations in data timeliness and completeness, showcase how we adapted these forecasts to support decisions of public health priority in rapidly-evolving situations, evaluate the impact of key model features on forecast skill, and demonstrate how to assess forecast skill in real-time before the ground truth is known. Conditioning the model on the most recent, but incomplete, data improved the forecast skill, emphasising the importance of developing strong quantitative models of surveillance system characteristics, such as ascertainment delay distributions. Forecast skill was highest when there were at least 10 reported cases per day, the circumstances in which authorities were most in need of forecasts to aid in planning and response.

## 1 Introduction

As of January 2021, Australia had effectively controlled COVID-19 transmission, experiencing prolonged intervals of local elimination throughout 2020 despite a steady influx of imported cases, which have been effectively managed by Australia’s hotel quarantine system [1]. Scenario modelling conducted in February 2020 informed Australia’s rapid and strong initial response [2], which was sufficient to control the import-driven first wave [3]. In particular, Australia’s definitive border measures, including mandatory 14-day hotel quarantine on arrival from overseas, was effective in limiting community exposure from infected international arrivals.

Nonetheless, in May 2020 a breakdown in infection prevention and control measures in the state of Victoria’s hotel quarantine system resulted in community exposure that developed into a second wave [4, 5]. COVID-19 (ancestral strain) case incidence peaked at 450 locally-acquired infections per day in the last week of July 2020 [5, 6]. This was substantially higher than the peak of 131 daily cases in the import-driven first wave. Despite the imposition of stringent inter-state movement restrictions, cross-border importation of cases into the state of New South Wales led to a long period of constrained community transmission in that state throughout 2020, kept in check by proactive public health case finding and quarantine without imposition of stringent social measures [7]. In Victoria, a combination of case finding, contact tracing, and prolonged restrictions on movement and gathering sizes were required to bring the second wave to an end in October 2020 [5, 8]. Subsequent border incursions in various states and territories were associated with small clusters and in some cases localised community transmission, prompting the imposition of strong public health responses supported by variably stringent social restrictions over days and weeks. The daily incidence of locally-acquired COVID-19 cases in each jurisdiction over the study period is shown in Figure 1.

**Figure 1:**
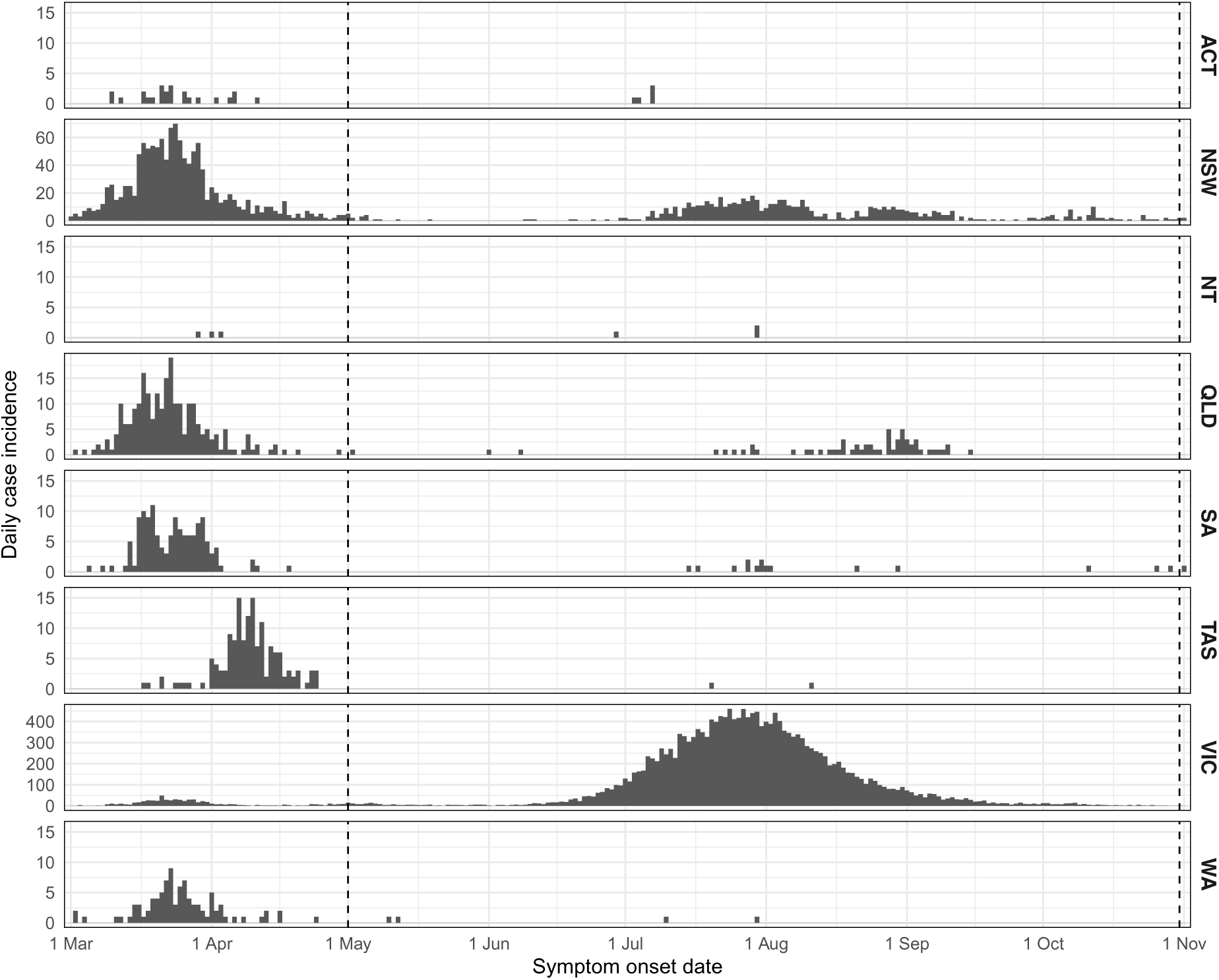
Daily incidence of locally-acquired COVID-19 cases for each jurisdiction, as reported after the end of the study period. Most jurisdictions experienced a first wave in March and April, comprised largely of imported cases. From June to October, Victoria experienced a large second wave and New South Wales experienced a prolonged low-amplitude small second wave. Vertical lines indicate the study period (1 May to 31 October). Note the different vertical scales for New South Wales and Victoria.

The public health response to the Victorian second wave and outbreaks in other Australian jurisdictions was informed by the results of real-time analytics and an ensemble forecast of COVID-19 activity for each Australian state and territory, presented to key government advisory committees in weekly situational reports. Similar analytics and forecasts have supported public health responses around the world [9–13]. Here we detail one of the forecasting models included in the ensemble forecast, which was adapted from Australian seasonal influenza forecasts [14–16] that we have deployed in near-real-time in collaboration with public health colleagues since 2015 [17, 18]. We evaluate the model’s performance by measuring forecast skill relative to an historical benchmark forecast over the study period (May to October 2020) and demonstrate how these outputs supported public health responses during that period. In addition to using the model to predict future rates of locally-acquired COVID-19 cases in each Australian jurisdiction, we demonstrate how our methods were adapted and improved in response to the needs of government, in order to most effectively support the public health decision-making process.

We generated forecasts by combining a stochastic SEEIIR compartment model with daily COVID-19 case counts through the use of a bootstrap particle filter [16]. The forecasting model incorporated real-time estimates of the effective reproduction number *R*_eff_ for the cohort of active cases (who may not be representative of the whole population), and the transmission potential (TP) averaged over the whole population (characterising the potential for wide-spread transmission) [7]. These latter estimates were informed by nationwide behavioural surveys and by mobility data from technology companies [7, 19]. We used the active-case *R*_eff_ to characterise local transmission at the time of forecast, and assumed that transmission from future active cases would gradually become more representative of the whole population. That is, local transmission in the forecasting model gradually shifted from the active-case *R*_eff_ to the whole-population transmission potential (TP) over the forecast horizon. We assess the impact of this assumption about active case heterogeneity by comparing the forecast skill against that of a separate suite of forecasts where local transmission was maintained at the current active-case *R*_eff_.

One of the most significant challenges in using predictive models to support public health decision-making in real time is understanding how much confidence to place in the predictions [20–22]. We address the question “When should we trust these forecasts?” by showing how forecast skill can be assessed in real-time *before the ground truth is known*.

## 2 Results

We first examine how well the forecasts characterised future case incidence during the onset of the second wave in Victoria (Section 2.1). Over this period, the timeliness and completeness of the near-real-time case data varied substantially, and we show how they impacted the forecast predictions.

We then demonstrate how we adapted the forecasts to support the gradual easing of restrictions in Victoria, as daily case incidence decreased in September and October but continued to persist at low levels (Section 2.2). Policy changes during this period were subject to reaching specific 14-day moving average case thresholds.

Finally, we present an analysis of the forecast performance across all Australian jurisdictions, in which we measure forecast skill against an historical benchmark forecast (Section 2.3). We identify the circumstances under which forecast skill was highest, evaluate the impact of key model features on forecast skill, and conclude by considering how to assess forecast skill in real-time.

### 2.1 Onset of the second wave in Victoria

Throughout May 2020, Victorian case incidence remained low and relatively stable (5.6 cases per day, on average). With such limited chains of local transmission, forecasts generated in May considered local extinction to be the most likely outcome and did not include the possibility of a large outbreak. These predictions were consistent with the findings of a subsequent genomic study, in which the month of May 2020 was characterised by near elimination of COVID-19 in Victoria [23]. However, a new cluster of local cases emerged at the end of May [23], and by late June more widespread epidemic activity was clearly established, with more than 60 cases per day (Figure 1).

Here we focus on the following key questions: “When did the forecasts first consider a large outbreak *was possible*?”; “When did the forecasts first consider a large outbreak *was likely*?”; and “How did the reported data available at each week influence these predictions?”. Recall that each forecast was conditioned on the expected case counts by symptom onset date, after accounting for delayed case ascertainment (see Sections 4.1–4.2 for methodological details).

#### 2.1.1 Likelihood of a large outbreak

The forecast generated on 24 June was the first to indicate that a large outbreak *might occur*. Forecasts generated on this date, and for the subsequent six weeks, are shown in Figure 2, and a qualitative summary of each forecast is provided in Table 1.

**Figure 2:**
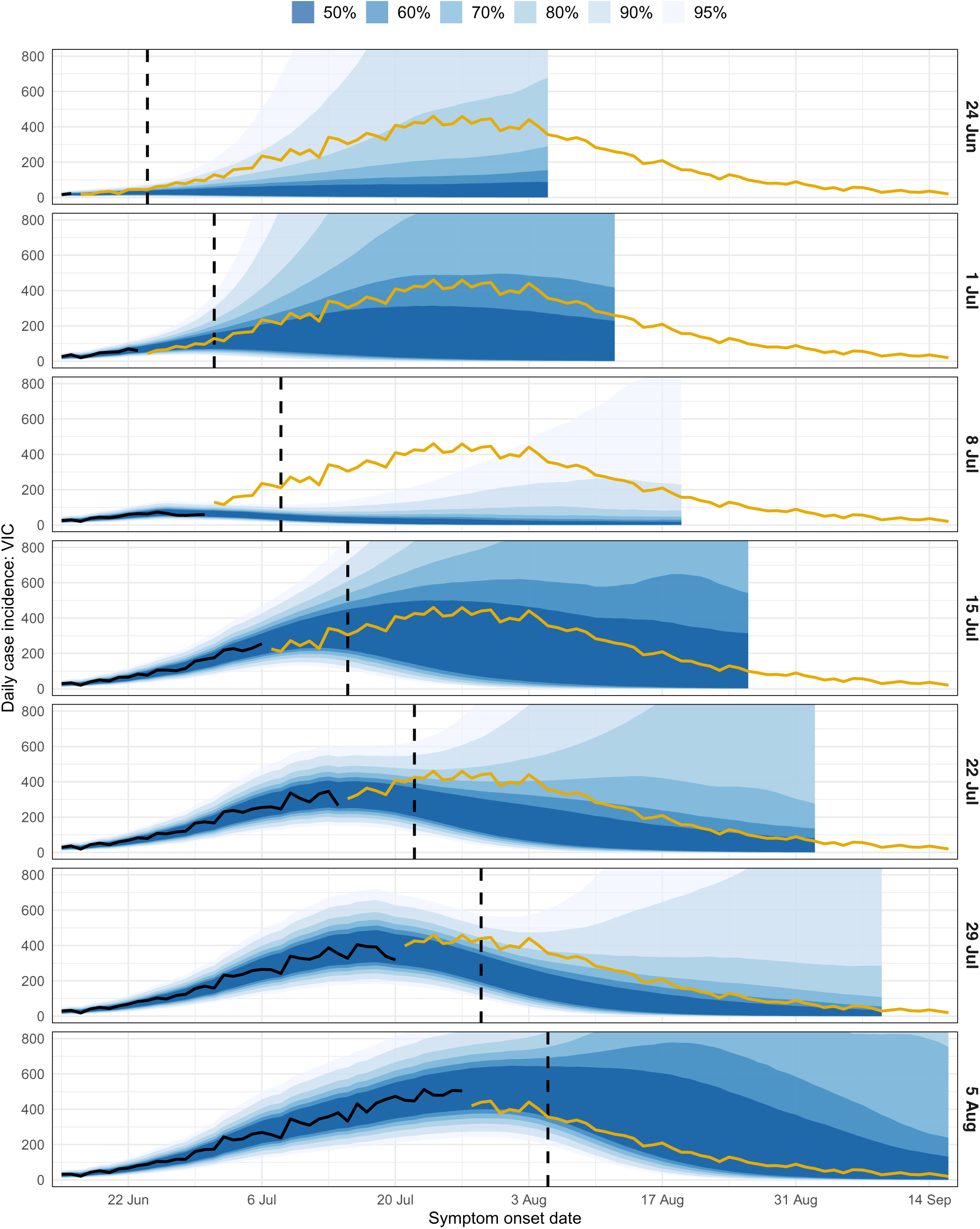
Forecasts of daily COVID-19 case incidence for Victoria, shown separately for each data date (right-hand axis labels). Each forecast is shown as a spread of credible intervals (blue shaded regions). Vertical dashed lines indicate the data date (i.e., when the forecast was generated). Black lines show the daily case counts at the time of forecasting, after accounting for delayed ascertainment, and dark yellow lines show the future case counts. Note that the daily case counts at the time of forecasting (black lines) end several days prior to the data date; see Section 4.2.2 for details.

**Table 1:** Qualitative summaries of the forecast credible intervals (CrIs) during the onset of the second wave, where “Date” is the date on which each forecast was generated.

| Date | Forecast characteristics |
| --- | --- |
| 24 June | 50% CrI is flat (no outbreak), 95% CrI includes possibility of large outbreak |
| 1 July | 50% CrI upper bound in good agreement with future case counts |
| 8 July | 95% CrI undershoots future case counts |
| 15 July | 50% CrI includes all of the future case counts |
| 22 July | 50% CrI undershoots future case counts after the peak |
| 29 July | 50% CrI undershoots future case counts after the peak |
| 5 August | 50% CrI includes all of the future case counts |

The forecast generated on 1 July was the first to demonstrate confidence that a substantial outbreak *would occur*. Its median trajectory exceeded 100 cases per day, the 50% credible interval (CrI) lower bound remained above 10 cases per day until mid-August, and the 50% CrI upper bound was in good agreement with the future case counts over the entire forecast horizon. As local transmission became more established, subsequent forecasts yielded increasingly confident and accurate predictions of the size and timing of this second wave. The 8 July forecast was a marked outlier from the other forecasts over this period, and we address this forecast separately in the next section.

Despite the 50% CrIs being reasonably narrow, the 95% CrIs remained very broad in all of these forecasts. The 95% CrI lower bounds decreased to zero cases per day roughly 2 weeks after the forecast date, and their upper bounds exceeded 1,000 cases per day. This diversity of epidemic trajectories (from the stochastic SEEIIR compartment model) illustrates the inherent uncertainty at the time of each forecast about the future course of this second wave, and what impact the imposed public health and social measures might have on local transmission over the forecast horizon.

#### 2.1.2 The impact of near-real-time data

A natural consequence of working with near-real-time surveillance data was the challenge of interpreting the reported numbers of locally-acquired cases for recent dates, given the delay between symptom onset and cases being ascertained and reported. Figure 3 shows the daily case counts reported at each data date, and the expected daily case counts after accounting for delayed ascertainment [7] (see Section 4.2 for methodological details). Ascertainment delays are evident in the data reported on and after 24 June (i.e., once there was an indication of established local transmission) and became more pronounced as local case incidence increased. The most recent data in each extract (dashed lines) systematically undershoot the corresponding case counts in subsequent data extracts.

**Figure 3:**
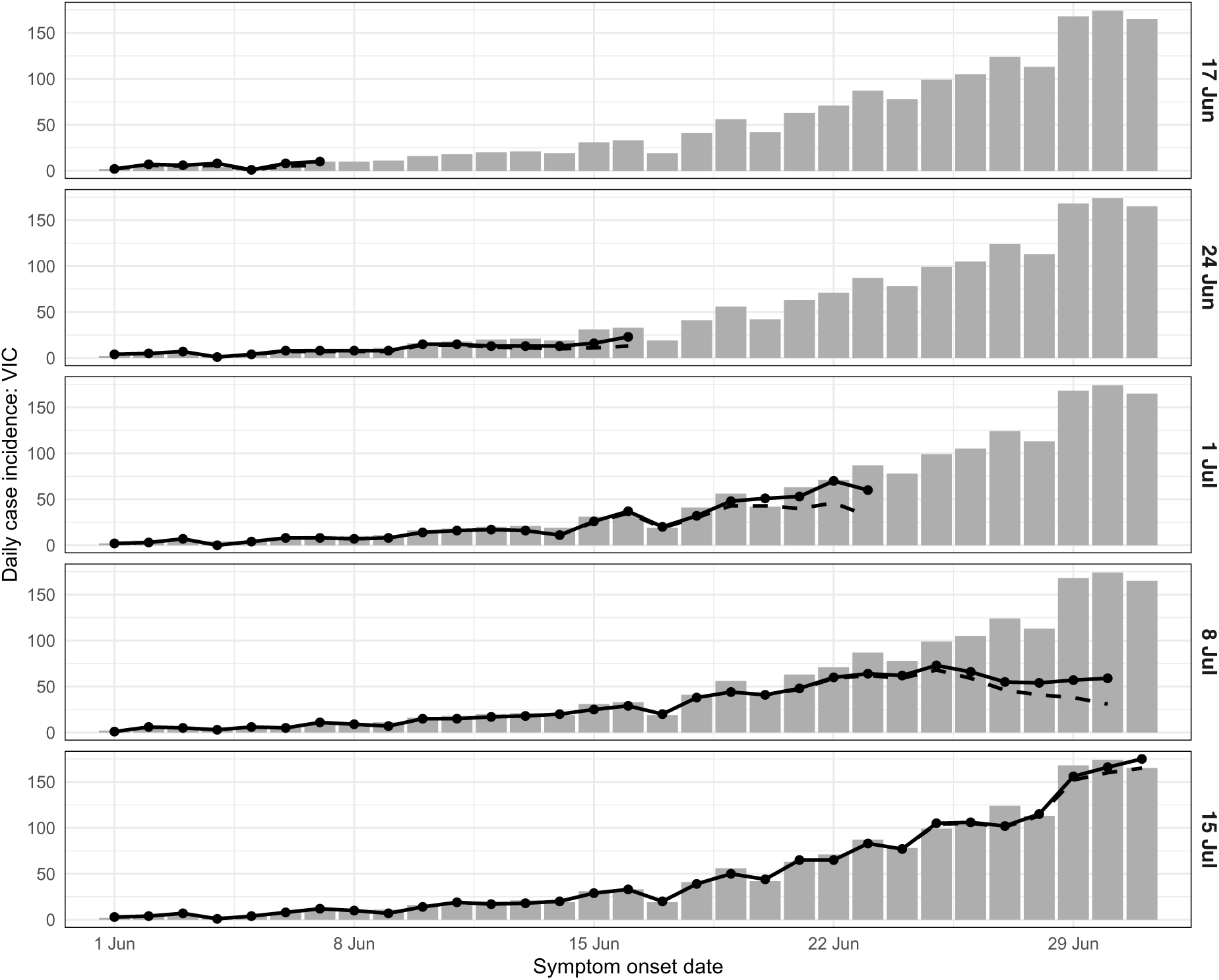
Daily incidence of COVID-19 cases for Victoria from mid-May until the end of June, shown separately for each data date (right-hand axis labels). Each data extract updated the entire time-series. Forecasts were conditioned on the expected case counts after accounting for delayed ascertainment (solid black lines); the reported case counts in the data extracts are also shown (dashed black lines). Updated case counts, as reported on 12 August, are shown as grey columns.

The forecast generated on 8 July is a particularly notable example of how data timeliness and completeness can impact model predictions. Its 50% CrI is much narrower than those of other forecasts and trends downward from the most recent observation (30 June), undershooting every future daily case count, even those before the data date itself. This forecast was influenced by an apparent decrease in daily cases reported over 25–30 June (Figure 3), even after applying the modelled correction for delays between symptom onset and case ascertainment (based on data from earlier data extracts), and might have revealed an opportunity for achieving elimination. However, case counts for these dates were substantially higher in subsequent data extracts. While there were a number of local clusters in Victoria around this time, including in multiple schools and public housing sites [24, 25], to the best of our knowledge this ascertainment delay was not linked to any specific cluster. A plausible explanation is that unprecedented case numbers impacted various aspects of the public health response (e.g., laboratory testing, contact tracing, reporting). The Victorian Government subsequently received additional support from Australian Defence Force personnel [26].

In contrast, the forecast generated on 5 August is in good agreement with the data, and its 50% CrI includes all of the future case numbers. Note the raw data available at the time of forecast suggested an epidemic in decline, but incidence remained near-maximal for the next two weeks and this was captured by the forecast.

These observations highlight the importance and the challenge of understanding the *meaning* and *limitations* of the available data in near-real-time. While correcting for delays (right censoring) is necessary, such corrections are not static through time. It is fundamentally difficult to estimate the completeness of these data in rapidly-evolving situations, such as during rapid epidemic growth, where bottlenecks and delays are not directly observable and are subject to change. Active communication with those responsible for the data is crucial to ensuring that the data and the resulting forecasts are interpreted appropriately, particularly when there are changes in surveillance, testing, and/or reporting.

### 2.2 Policy context: easing restrictions in Victoria

In response to the imposed restrictions (“stage 3” and “stage 4”), daily case numbers in Victoria peaked in August and steadily decreased thereafter [5, 7]. By early September the Victorian government had announced a plan to gradually ease these restrictions over a period spanning mid-September to late November, conditional on achieving specific case thresholds over 14-day windows ahead of each planned date for policy changes [27]. However, similar to the months of May and June, local cases remained at low but persistent levels in the second half of September and into October (see Figure 4). The city of Melbourne had been under intense mobility and gathering restrictions (so-called “lockdown”) since 7 July, and the growing need to ease these restrictions had to be carefully balanced against the risk of resurgence. We used our forecasts to predict the probability that the 14-day moving average for incident cases would achieve these specific thresholds at each day of the forecasting period (see Section 4.5 for methodological details). Three to four weeks in advance of each threshold being reached, the forecasts predicted a greater than 50% chance of reaching the threshold by the date it was reached (see Table 2 and Figure 5). The one exception was the 10-case threshold, which was reached on 9 October. In the weeks prior to this date, daily case counts fluctuated around 10 cases per day (see Figure 4). Three weeks prior to this threshold being reached, the forecasts predicted only a 23% chance of reaching this threshold by 9 October. By the following week — two weeks in advance — the forecasts predicted a 68% chance of reaching this threshold by 9 October.

**Figure 4:**
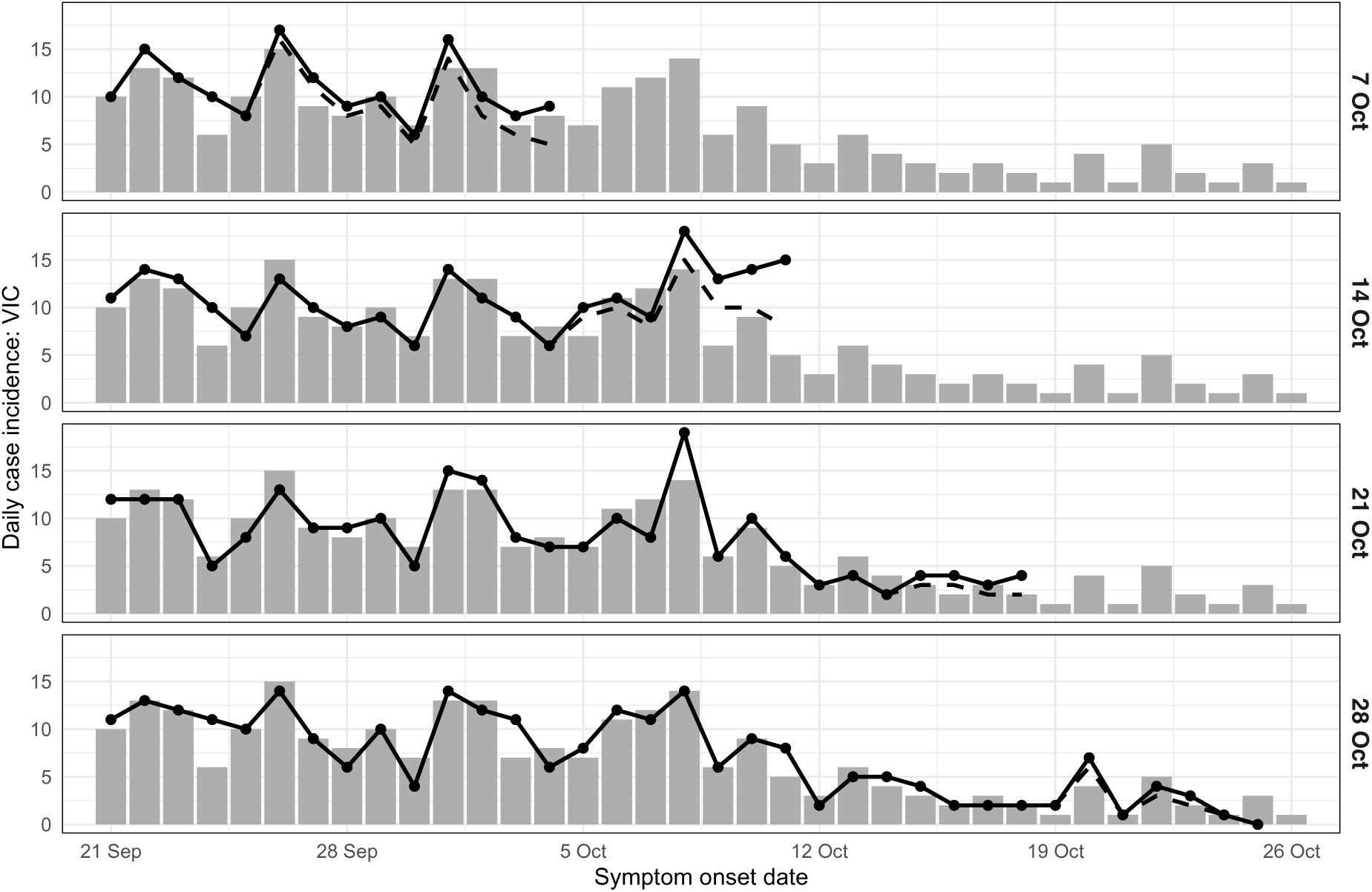
Daily incidence of COVID-19 cases for Victoria in October, shown separately for each data date (right-hand axis labels). Forecasts were conditioned on the expected case counts after accounting for delayed ascertainment (solid black lines); the reported case counts in the data extracts are also shown (dashed black lines). Updated case counts, as reported after the study period, are shown as grey columns.

**Figure 5:**
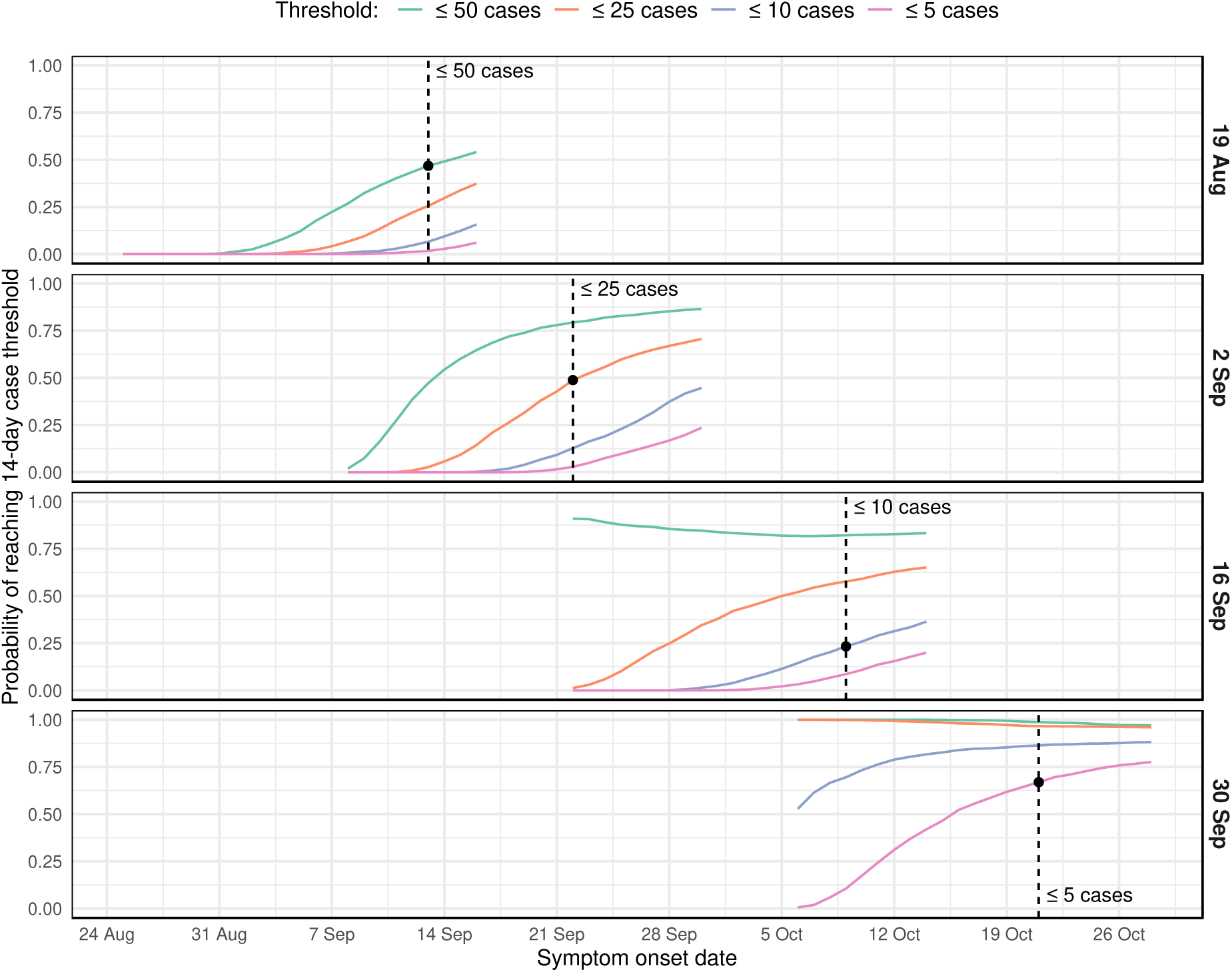
Time series showing the projected probability of reaching each of the 14-day moving average case thresholds, shown separately for each data date (right-hand axis labels). Vertical dashed lines show when each threshold was first reached (with respect to symptom onset date, not reporting date). The black points show the projected probability of having achieved each threshold on the day that it was reached.

**Table 2:** Forecast predictions concerning the 14-day moving average case thresholds. For each threshold, this table shows the date when that threshold was first reached, and the following forecast quantities for that date: (a) the forecast data date; (b) the projected probability of reaching the threshold by the date when it was first reached; and (c) the forecast 50% credible interval for daily case incidence on that date.

| Threshold | First reached | Data date | Probability | 50% CrI |
| --- | --- | --- | --- | --- |
| $\leq 50$ cases | 13 Sep | 19 Aug | 46.8% | 7–94 |
|  |  | 26 Aug | 63.5% | 8–44 |
| $\leq 25$ cases | 22 Sep | 26 Aug | 64.1% | 1–24 |
| $\leq 10$ cases | 9 Oct | 23 Sep | 68.2% | 1–8 |
| $\leq 5$ cases | 21 Oct | 23 Sep | 72.7% | 0–4 |

### 2.3 Forecast skill

Having described the behaviour and qualitative performance of our forecasts during periods of increasing, decreasing, and low levels of local transmission, we now consider their performance more precisely, through skill scores. We measured forecast skill relative to an historical benchmark, described in Section 4.4, which assumed that future case counts would follow the same distribution as the case counts observed to date. This provides a quantitative performance evaluation and helps to address a critical question when using forecasts to support public health decision-making: “When should we trust these forecasts?”.

#### 2.3.1 Forecast uncertainty

To quantify forecast uncertainty we used *forecast sharpness,* a measure of the breadth of forecast outcomes (e.g., a sharpness of zero means there is a single possible outcome). For jurisdictions that reported substantial COVID-19 activity, as the lead time increased (i.e., looking further into the future) the forecasts exhibited little increase in uncertainty, or even grew more certain (Section B.1). This trend reflects, in part, that as local epidemic activity increased there was more information available to inform the forecasts and constrain the range of plausible outcomes. During the second wave in Victoria, strong public health measures and social restrictions markedly reduced the TP, and this caused the forecasts to become sharper at longer lead times (Figure S2). In contrast, assuming that transmission would be sustained at the current active-case *R*_eff_ caused the forecasts to become more uncertain at longer lead times (Figure S3). For jurisdictions that primarily reported zero cases on a given day, the forecast trajectories tended to local extinction, and forecast sharpness was close to zero (i.e., minimal uncertainty) for all lead times.

#### 2.3.2 Forecast bias and case heterogeneity

The forecasts tended to under-estimate future case counts (negative bias, see Section B.2). For jurisdictions which had achieved local elimination, such as South Australia in May, this reflects a deliberate model feature: our careful separation of the *potential* for reintroduction from the *consequence* of reintroduction. Without evidence of reintroduction into these jurisdictions, the forecasts did not admit the possibility of subsequent epidemic activity (see Section 4.3) and predicted there would be zero reported cases each day. On the rare occasions where a jurisdiction subsequently reported one or two isolated cases (see Figure 1) this resulted in a negative bias (under-estimation).

For Victoria, which experienced substantial COVID-19 activity during the study period, model assumptions about local transmission contributed to the underestimation of future case counts. Recall that the model used the active-case *R*_eff_ to characterise local transmission at the time of forecast, with transmission reverting to the transmission potential (TP) over time (see Section 4.2 for details; example *R*_eff_ trajectories are shown in Section B.4). However the second wave in Victoria was dominated by localised outbreaks in health and aged care settings, and other essential services, where public health and social measures to constrain transmission had less impact than in the general population [5]. Over a 5-month period, the *R*_eff_ was systematically greater than the TP, as described and evaluated in Golding et al. [7]. Accordingly, as shown in Figure 2, the majority of the Victorian forecasts somewhat under-estimated future case counts. Despite this negative bias, the model consistently out-performed forecasts where local transmission was instead maintained at the current active-case *R*_eff_ (see Section B.3.1). Noting that *R*_eff_ was consistently higher than the TP, this highlights the importance (and challenge) of accounting for heterogeneity of transmission in diverse settings and population sub-groups.

#### 2.3.3 Forecast skill and the historical benchmark

Of the five jurisdictions that primarily reported zero cases on a given day, the forecasts consistently out-performed the historical benchmark for all lead times in the Australian Capital Territory and Western Australia, and out-performed the historical benchmark for 1-week lead times in South Australia and Tasmania. The historical benchmark outperformed the forecasts for all lead times in the Northern Territory, where the past case counts were extremely good predictors of the future case counts. For Queensland, which saw a sustained period of 1–5 cases per day (July to September 2020) the forecasts out-performed the historical benchmark for lead times of up to 2 weeks. For longer lead times the forecasts tended to include the possibility of a sustained increase in cases, which was inconsistent with the ground truth and reduced their skill relative to the historical benchmark.

For jurisdictions that experienced substantial COVID-19 activity (New South Wales, Victoria) the forecasts exhibited improved performance relative to the historical benchmark for shorter lead times (Section B.3.3). For Victoria, which experienced the largest local outbreak, the forecasts substantially out-performed the historical benchmark for lead times of up to 4 weeks (Section B.3.4).

Forecast skill across all jurisdictions was highest when daily case numbers were zero or at least ten (Section B.3.1). The forecasts also out-performed the historical benchmark when there were 1–9 daily cases, but to a lesser degree. Across all jurisdictions, 1–9 daily cases comprised 18% of the daily observations; 4% in the state of Victoria, the other 14% in jurisdictions that did not experience sustained epidemic activity. These are the circumstances under which there is great uncertainty as to whether local transmission will become established and drive exponential growth in case numbers. The model forecasts began to include the possibility of a large outbreak in these circumstances (e.g., see the Victorian forecast for 24 June in Figure 2) and many trajectories substantially exceeded the future case counts, since enacted control measures regularly curtailed transmission.

In contrast, the historical benchmark predictions were defined by the daily case counts reported to date and yielded much narrower predictions than the forecast model in these circumstances. The majority of the 1–9 daily case observations occurred in jurisdictions that did not experience sustained epidemic activity, for which the past observations were good predictors of the future case counts, and the historical benchmark performed well. For Victoria, the historical benchmark predictions substantially underestimated the future case counts in the second wave, and did not include the ground truth as a possibility.

Of course, when considering the model forecasts as a risk evaluation tool, their ability to include the potential for large outbreaks in their projections is a major advantage over the historical benchmark, as is their ability to predict peak timing and size.

#### 2.3.4 Forecasting in near-real-time

We highlighted some challenges of using near-real-time data in Section 2.1. By accounting for delayed ascertainment in the reported case counts (depicted in Figures 3 and 4) we extracted additional information that, on average, increased the forecast skill (Section B.3.2), even though it slightly decreased the forecast skill for days where 1–9 cases were reported (Section B.3.1). Forecast performance was particularly sensitive to the imputed case counts when the reported case counts were low and a difference of, say, 1 case per day represented a large relative change (see Figures 3 and 4 for examples).

When considering forecast skill as a function of the number of cases ultimately recorded for each day, as above, the results can only be evaluated in retrospect, since these numbers are unknown when the forecast is made. Accordingly, we also considered forecast skill as a function of the number of daily cases *predicted by the forecast model* (Section B.3.5), which may provide assistance when interpreting the forecast outputs in near-real-time. Our intent was to address the question “What does this forecast mean?” before the ground truth is known.

The skill scores were similar when aggregated by the number of cases ultimately reported for each day or by the median number of predicted case numbers, particularly when there were at least 5 cases per day (Figure S17). This indicates that in decision-making contexts (i.e., when future case counts are unknown) we can still obtain a reasonable estimate of forecast skill and assess forecast reliability in real-time. These estimates are particularly reliable when local transmission is established.

## 3 Discussion

### 3.1 Principal findings

Forecasting infectious disease activity is a fundamentally difficult problem [28], and even more so in unprecedented circumstances and in the face of regularly updated public-health responses, public perceptions of risk, and changes in behaviour [29–31]. Despite these challenges, we were able to adapt our existing seasonal influenza forecasting methods [14–18] to COVID-19 transmission in Australia, and provide the resulting forecasts to public health authorities to support the Australian COVID-19 response. We tailored our analyses in near-real-time to support public health decision-making in rapidly-evolving situations. This included providing public health authorities with estimated future epidemic activity, timelines for achieving case thresholds that would trigger easing of restrictions, and risk assessments based on repeated incursions of new chains of transmission in primarily infection-free jurisdictions.

Our forecasts exhibited the greatest skill in the state of Victoria, which experienced the overwhelming majority (96%) of locally-acquired cases reported in Australia over the study period, May–October 2020. Forecast skill was highest when there were at least 10 reported cases on a given day, the circumstances in which authorities were most in need of forecasts to aid in planning and response.

In considering several alternate forecast models, forecast skill was higher when we assumed that the active-case *R*_eff_ characterised local transmission at the time of forecast and that, over the forecast horizon, local transmission would trend back towards the whole-population transmission potential (whether *R*_eff_ was lower or higher than the TP). This improvement in skill was driven by characteristics of both the Victorian and New South Wales epidemics. In Victoria, while *R*_eff_ remained consistently higher than the transmission potential, it decreased steadily over time [7]. In New South Wales, the public health response to incursions repeatedly drew *R*_eff_ below the TP [7]. Although our model overestimated the rate at which local transmission would trend back towards the TP, it consistently out-performed forecasts generated under the assumption that local transmission would be sustained at the current active-case *R*_eff_.

Forecast skill was also higher when the model was conditioned on the most recent data, even though these data were known to be incomplete. By imputing symptom onset dates and estimating the delay between symptom onset and case ascertainment [7], we obtained additional information to that available in the reported time-series data, improving forecast skill. However, when the reported number of daily cases were low, forecast performance was particularly sensitive to the imputed case counts and there were occasions where forecast skill was higher when the model was not conditioned on the most recent data.

### 3.2 Study strengths

Australian jurisdictions maintained high testing levels over the study period. The proportion of infected persons that were identified as cases was likely to be both very high, and to remain relatively constant over the study period. These cases include asymptomatic individuals who were identified through large-scale and systematically exhaustive contact tracing programs [5]. In comparison, in recent influenza seasons the probability that a person with influenza-like illness (ILI) symptoms would seek healthcare and have a specimen collected for testing (as estimated from Flutracking [32, 33] survey participants) was around 3–8% [16]. But during the study period, 50–75% of Flutracking participants with ILI symptoms reported having a COVID-19 test (Section B.5), suggesting high case ascertainment. The end of the second wave in Victoria provides yet further evidence of high case ascertainment. As the 14-day case thresholds for easing restrictions were achieved, there were fewer and fewer unlinked (“mystery”) cases reported. For example, the Victorian 19 October update reported an average of 7.7 cases per day in the past 14 days and a total of just 15 unlinked cases [34], while the 26 October update reported an average of 3.6 cases per day in the past 14 days and a total of just 7 unlinked cases [35]. Elimination was achieved before the end of the study period, and so unidentified cases, if present, did not cause sustained chains of transmission. This is consistent with the TP, which was estimated to be less than one in Victoria from August to October 2020 [7].

As described above, we had access to detailed line-listed data, and were able to estimate the delay distribution for the time between symptom onset and case ascertainment, and to impute the symptom onset date for cases where this had not yet been reported [7].

We were able to incorporate the impact of public health interventions via near-real-time estimates of the effective reproduction number (*R*_eff_) and the whole-population transmission potential (TP), which were informed by nationwide behavioural surveys, population mobility data, and times from symptom onset to case detection [7, 19]. Upon reintroduction of cases in jurisdictions that had achieved local elimination, estimates of the transmission potential allowed us to produce forecasts that were informed by local mobility and behaviour, before reliable estimates of the effective reproduction number were available.

Likewise, we were able to adapt the forecast outputs to support policy decisions in near-real-time for individual jurisdictions without incorporating additional effects into the underlying simulation model. This meant that we did not need to explicitly model control measures that were imposed and relaxed in each jurisdiction — such an effort would have been extremely complex and it would be impossible to account for decisions that occur within each forecast horizon and their impact on transmission dynamics. The fact that our forecasts exhibited high forecast skill, particularly during periods of sustained local transmission, demonstrates that increasing model complexity is not always necessary or appropriate [12, 22, 36, 37].

### 3.3 Study limitations

Australia had experienced limited COVID-19 activity prior to the study period, and so it was reasonable to assume that the entire population was susceptible to infection. Subsequent waves of local infection, as experienced in Australia after the study period, have resulted in a non-negligible proportion of the population having some degree of natural immunity. In these circumstances, the forecasts would become more sensitive to assumptions about case ascertainment. COVID-19 vaccination in Australia began in February 2021 (i.e., after the study period) and has mitigated the impact of subsequent circulation of the Delta and Omicron variants in 2021 and 2022. By February 2022, more than 94% of people over the age of 16 were fully vaccinated [38]. We have continued to refine and extend the methods presented here to account for changes in case ascertainment, vaccination coverage, booster vaccinations, and waning immunity, and these refinements will be described elsewhere.

Our forecast model only incorporated population heterogeneity by allowing local transmission to trend from the active-case *R*_eff_ to the whole-population transmission potential (TP) over the forecast horizon. This limited granularity was appropriate for the purposes to which the forecasts were applied, but meant that the forecasts were not suitable for addressing more nuanced questions about, for example, targeted (geographic or sociodemographic) interventions to mitigate localised clusters [39].

### 3.4 Meaning and implications

The first wave of COVID-19 in Australia was primarily driven by returning travellers who had acquired their infection overseas. In contrast, the second wave in Victoria was driven by local transmission. Cases in the second wave were also over-represented in essential services that involved frequent and intense contact, such as residential aged care facilities, healthcare, manufacturing, and meat processing [5], and so were less able to reduce their contacts (and so transmission opportunities) than the general population. Accordingly, during the second wave in Victoria the active-case *R*_eff_ was systematically higher than the whole-population transmission potential [7], even though both decreased over time in response to public health interventions. Forecast bias was compounded by this disparity between *R*_eff_ and transmission potential.

However, since the active-case *R*_eff_ did decrease over time, forecast skill was highest when, over the forecast horizon, local transmission trended back towards the whole-population transmission potential. It is unclear how future trends in local transmission could be better modelled or predicted, highlighting fundamental challenges in regards to (a) understanding the transmission characteristics of persons who are currently infectious; and (b) predicting how the transmission characteristics of infectious persons will change into the future, particularly when case numbers are low. These are intrinsic sources of uncertainty for infectious diseases forecasting, and while they may conceivably be reduced, they cannot be eliminated.

There is an acknowledged need for effective and on-going collaborations between modellers and public health practitioners in order to take full advantage of modelling tools such as epidemic forecasts to support public health policy and decision-making [40–42]. The forecasts presented here were adapted from our Australian seasonal influenza forecasts [14–16], which we have deployed in near-real-time in collaboration with public health colleagues since 2015 [17, 18]. A number of modifications were required to adapt these forecasts to COVID-19 transmission in Australia, but developing and applying these methods to seasonal influenza activity in previous years has provided us with valuable experience in adapting to unprecedented circumstances, working with incomplete and evolving data, responding to public health needs, and effectively communicating forecast outputs to public health decision-makers.

## 4 Materials and methods

### 4.1 COVID-19 surveillance data

We used line-lists of reported cases for each Australian state and territory, which were extracted from the National Notifiable Disease Surveillance System (NNDSS) on a weekly basis. The line-lists contained the date when the individual first reported exhibiting symptoms, the date when the case notification was received by the jurisdictional health department, and whether the infection was acquired locally or overseas.

We used a time-varying delay distribution to characterise the duration between symptom onset and case notification, which was estimated from the reporting delays observed during the study period [7]. Symptom onset dates were imputed for cases where this was not (yet) reported.

### 4.2 Forecasting COVID-19 case numbers in the community

We used a discrete-time stochastic SEEIIR model to characterise infection in each Australian jurisdiction. Let *S*(*t*) represent the number of *susceptible* individuals, *E*_1_(*t*)+*E*_2_(*t*) represent the number of *exposed* individuals, *I*_1_(*t*) + *I*_2_(*t*) represent the number of *infectious* individuals, and *R*(*t*) the number of *removed* individuals, at time *t*. Symptom onset was assumed to coincide with the transition from *I*_1_ to *I*_2_. We used real-time estimates of the effective reproduction number *R*_eff_ [7], which were informed by nationwide behavioural surveys, mobility data from technology companies, and times from symptom onset to case detection [7, 19], and sampled a unique trajectory 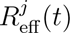 for each particle *j*. In order to initialise the first epidemic wave in each jurisdiction, it was assumed that 10 exposures were introduced into the *E*_1_ compartment at time *τ*, to be inferred, giving initial conditions:

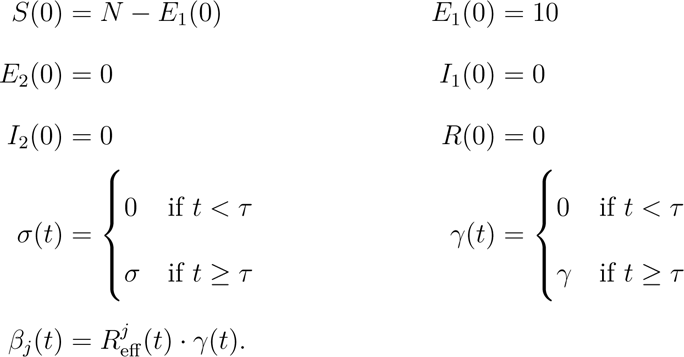

The number of individuals who leave each compartment on each time-step Δ*t* follows a binomial distribution (see Table 3 for parameter definitions):

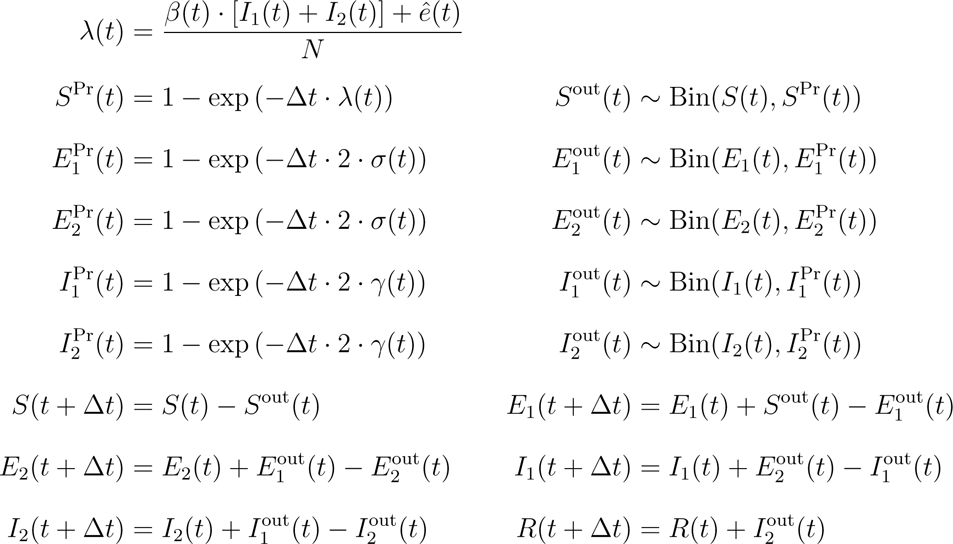

**Table 3:**
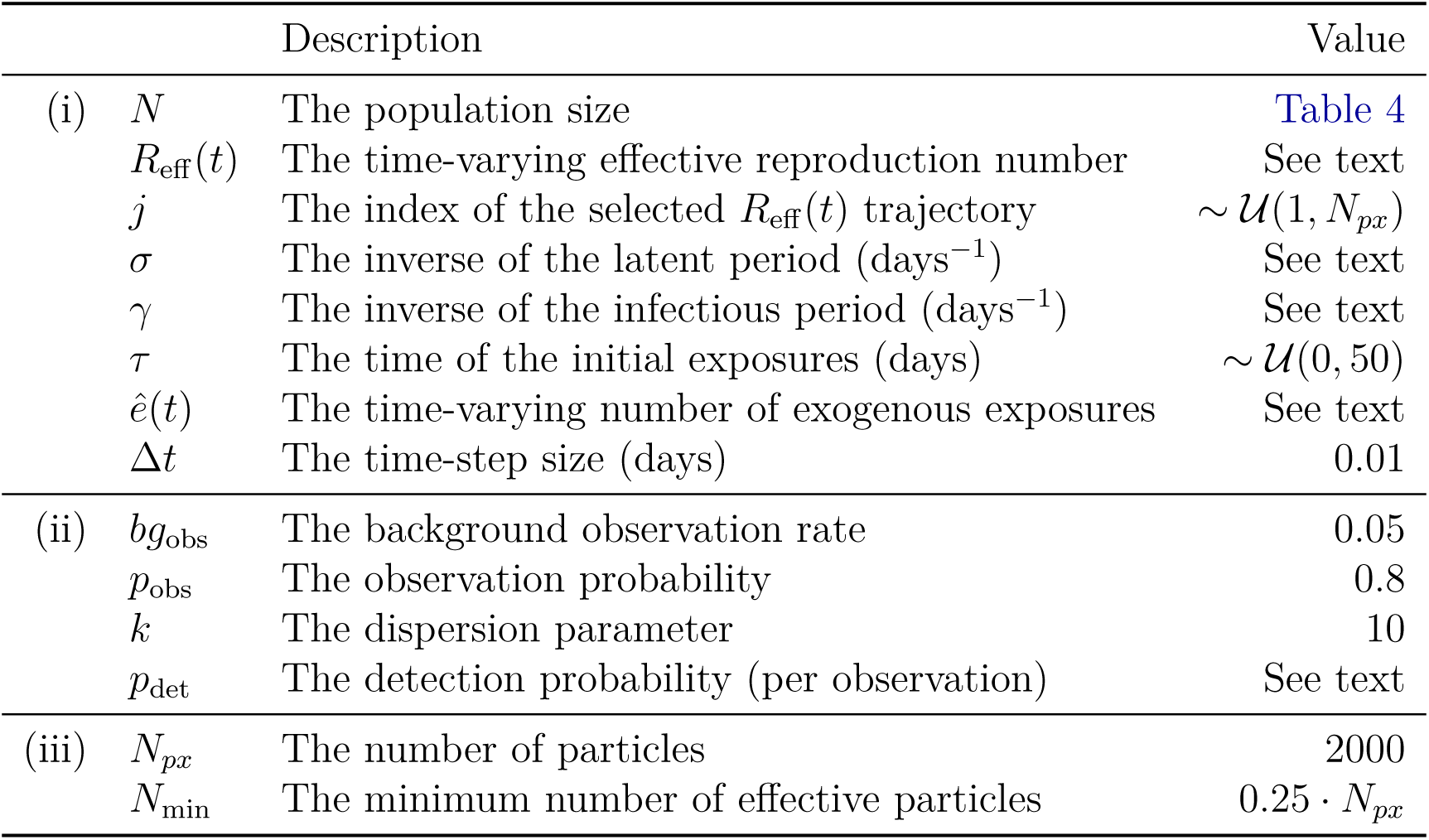
Parameter values for (i) the transmission model; (ii) the observation model; and (iii) the bootstrap particle filter.

We allowed for the introduction of exogenous exposures, which may be acquired by individuals who, e.g., visit an external location where infectious individuals are present, or encounter an infectious individual from an external location who has travelled to this location. The expected number of exogenous exposures, *ê*(*t*), is time-varying and independent trajectories can be sampled for each particle.

We modelled the relationship between model incidence and the observed daily COVID-19 case incidence (*y_t_*) using a negative binomial distribution with dispersion parameter *k*, since the data were non-negative integer counts and were over-dispersed when compared to a Poisson distribution. Let X(*t*) represent the state of the dynamic process *and* particle filter particles at time *t*, and *x_t_* represent a realisation; *x_t_* = (*s_t_, e*_1*t*_*, e*_2*t*_*, i*_1*t*_*, i*_2*t*_*, r_t_, σ_t_, γ_t_, j_t_, ê_t_*). We assumed that cases were observed (i.e., were reported as a notifiable case) with probability *p*_obs_ when they transitioned from *I*_1_ to *I*_2_. To account for incomplete observations due to, e.g., delays in testing and reporting for the most recent observations, we fitted a time-to-detection distribution and calculated the detection probability (*p*_det_) for each observation at each time *t* [7]. In order to improve the stability of the particle filter for very low (or zero) daily incidence, we also allowed for the possibility of a very small number of observed cases that were *not* directly a result of the community-level epidemic dynamics (*bg*_obs_). Accordingly, we defined the likelihood ℒ(*y_t_ | x_t_*) of obtaining the observation *y_t_* from the particle *x_t_* as:

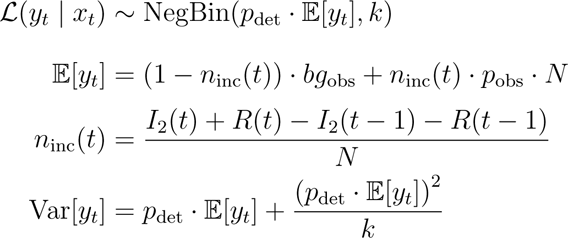

The value of the dispersion parameter *k* was chosen so that when zero individuals enter the *I*_2_ compartment over one day, the 50% credible interval has an upper bound of zero observed cases and the 95% credible interval has an upper bound of one observed case.

To generate projected case counts at each day, we used a bootstrap particle filter with post-regularisation [43] and a “deterministic” resampling method [44], as previously described in the context of our Australian seasonal influenza forecasts [14–18]. We constructed a simulated trajectory 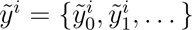 from each particle *i* in the ensemble by drawing a random sample 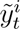 at each day *t*:

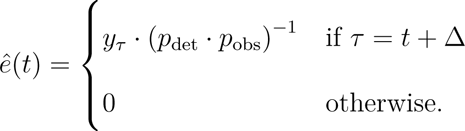

#### 4.2.1 Parameters and model prior distributions

Model and particle filter parameters are described in Table 3. Since Australia is one of the most urbanised countries in the world, for each jurisdiction we used capital city residential populations (including the entire metropolitan region, as listed in Table 4) in lieu of the residential population of each jurisdiction as a whole. Parameters *σ* and *γ* were sampled from a multivariate log-normal distribution that was defined to be consistent with a generation interval distribution with mean=4.7 and SD=2.9 [7, 45], and we sampled independent 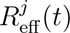 trajectories for each particle. The observation probability *p*_obs_ was fixed at 0.8, assuming that 80% of infections would be detected [46].

**Table 4:**
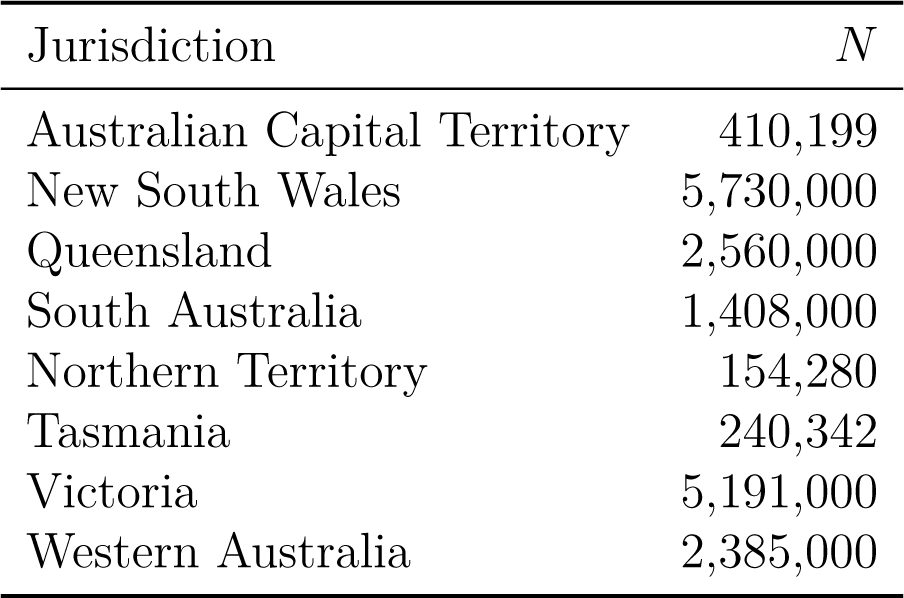
The population sizes used for each jurisdiction.

#### 4.2.2 Competing models

We evaluated four competing models, by considering all combinations of:

1. Assuming that local transmission would trend back towards the whole-population transmission potential over the forecast horizon (“Trend to TP”), or that local transmission would instead be sustained at the active-case *R*_eff_ distribution over the forecast horizon (“Fixed Reff”); and
2. Conditioning on all daily case counts where the detection probability *p*_det_ *≥* 0.5, or only conditioning on all daily case counts where *p*_det_ *≥* 0.95 (in order to reduce the influence of imputed symptom onset dates, and cases potentially not yet reported).

The labels used to identify each of these four models are listed in Table 5.

**Table 5:** The four competing forecast models.

| | Trend to TP | Fixed $R_{\text{eff}}$ |
| --- | --- | --- |
| $p_{\text{det}} \geq 0.5$ | Default | Fixed Reff |
| $p_{\text{det}} \geq 0.95$ | Minimal imputation | Minimal imputation, Fixed Reff |

### 4.3 Accounting for observed reintroduction

We deliberately treated the *consequence* of observed reintroduction separately from the *potential* for reintroduction. Here our interest lies in introducing new exposures to reflect a small number of reported cases in a jurisdiction after a sufficiently long period of no reported cases in that jurisdiction. This captures importation events where the particle ensemble contains few, if any, particles with any exposed or infectious individuals, and is fundamentally different from predicting the probability of a future reintroduction into a jurisdiction that currently has no active COVID-19 cases. We can realise this by defining exogenous exposure trajectories that are non-zero for a short period prior to the observed reintroduction.

For a jurisdiction that reports one or more COVID-19 cases at time *τ* (*y_τ_*) after a prolonged period of no cases (e.g., 28 days or longer), we assume that the initial exposure(s) occurred Δ = 5 days prior to the case(s) being reported, and we account for the time-to-detection distribution (*p*_det_) and the observation probability (*p*_obs_):

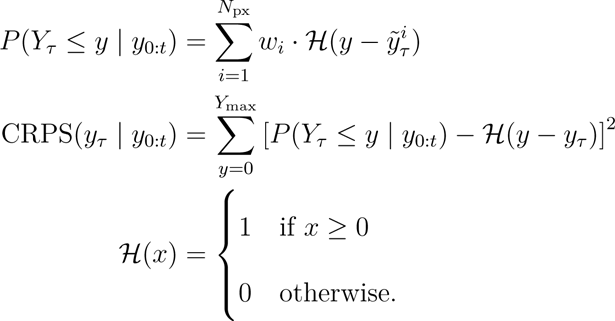

We deliberately selected this approach instead of keeping *ê*(*t*) fixed at some positive value *ε*, so that the forecasts for jurisdictions that had achieved local elimination did not suggest there would be subsequent epidemic activity without evidence of reintroduction. This decision was motivated by Australia’s international and domestic movement restrictions, which were effective in limiting community exposure from infected international arrivals and from travel between jurisdictions. The daily probability of an importation event was deemed to be sufficiently low that it would only serve to inflate the widest credible intervals in jurisdictions that had achieved elimination.

### 4.4 Forecast targets and skill

The forecast targets were the daily case counts (indexed by symptom onset date) for the 7 days prior to the data date (“back-cast”), for the data date itself (“now-cast”), and for the 28 days after the data date (“forecast”). Forecast performance was measured using Continuous Ranked Probability Scores (CRPS [47]) for each observation *y_τ_* of COVID-19 cases on day *τ*, given observations *y*_0:*t*_ for days 0 *… t* (inclusive):

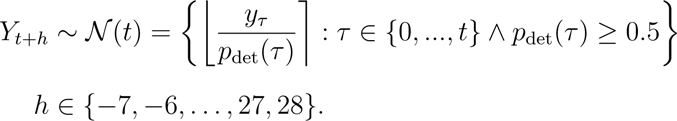

where ℋ denotes the Heaviside step function.

We compared the performance of each model ℳ to that of an historical benchmark forecast 𝒩, whose probability mass function 𝒩(*t*) for the data date *t* was the distribution of the daily case counts reported in the jurisdiction at the time of forecast, accounting for the detection probability *p*_det_(*τ*) for each observation *y_τ_* :

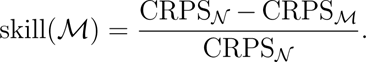

We measured the performance of each model ℳ using CRPS skill scores [48]:

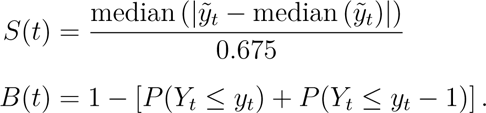

We also measured forecast sharpness *S*(*t*) and forecast bias *B*(*t*) as per Funk et al. [49]:

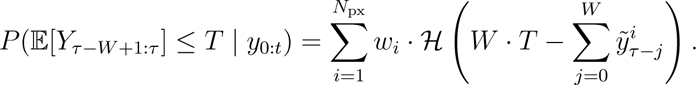

### 4.5 Meeting case-number thresholds that trigger policy changes

We have already defined the probability that the case counts on day *τ* will not exceed some threshold *T* : *P* (*Y_τ_ ≤ T | y*_0:*t*_). We can build on this definition to obtain the probability that the average daily case count over some window *W* up to, and including, day *τ* does not exceed this threshold *T* :

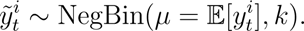

This was used to predict the probability of achieving each of the target case thresholds for relaxing measures in the second wave in Victoria, for each day in the forecast horizon.

## Data Availability

All of the simulation code and forecast outputs used to generate the results in this manuscript and the supplementary materials are provided in a public repository (https://gitlab.unimelb.edu.au/rgmoss/aus-2020-covid-forecasts). The input data are provided in a separate dataset (doi:10.26188/19315055).

https://doi.org/10.26188/19315055

## 5. Acknowledgements

National Notifiable Disease Surveillance System data on COVID-19 were provided by the Office of Health Protection and Response, Australian Government Department of Health and Aged Care, on behalf of the Communicable Diseases Network Australia (CDNA). We thank public health staff from incident emergency operations centres in state and territory health departments, and the Australian Government Department of Health and Aged Care, along with state and territory public health laboratories. We thank members of CDNA for their feedback and perspectives on the study results. We thank Craig Dalton, Sandra Carlson, and the Flutracking team for sharing their insights and Flutracking survey data throughout the study. We thank the Flutracking participants who made the time each week to complete the surveys and contribute to COVID-19 surveillance.

This research was supported by use of the Nectar Research Cloud, a collaborative Australian research platform supported by the NCRIS-funded Australian Research Data Commons (ARDC), and by the University of Melbourne’s Research Computing Services.

This work was directly funded by the Australian Government Department of Health. Additional support was provided by the National Health and Medical Research Council of Australia through its Centres of Research Excellence (SPECTRUM, GNT1170960) and Investigator Grant Schemes (JMcV Principal Research Fellowship, GNT1117140).

## 6 Ethics statement

The study was undertaken as urgent public health action to support Australia’s COVID-19 pandemic response. The study used data from the Australian National Notifiable Disease Surveillance System (NNDSS) provided to the Australian Government Department of Health and Aged Care under the National Health Security Agreement for the purposes of national communicable disease surveillance. Data from the NNDSS were supplied after de-identification to the investigator team for the purposes of provision of epidemiological advice to government. Contractual obligations established strict data protection protocols agreed between the University of Melbourne and sub-contractors and the Australian Government Department of Health and Aged Care, with oversight and approval for use in supporting Australia’s pandemic response and for publication provided by the data custodians represented by the Communicable Diseases Network of Australia. The ethics of the use of these data for these purposes, including publication, was agreed by the Department of Health and Aged Care with the Communicable Diseases Network of Australia.

## A Supporting materials

All of the simulation code and forecast outputs used to generate the results in this manuscript and the supplementary materials are provided in a public git repository: https://gitlab.unimelb.edu.au/rgmoss/aus-2020-covid-forecasts

See the provided README.md file for detailed instructions.

The input data for each data date (i.e., the daily COVID-19 case counts and R_eff_ trajectories) are provided in a separate dataset (https://doi.org/10.26188/19315055).

## B Supplementary figures and results

### B.1 Forecast sharpness

**Figure S1:**
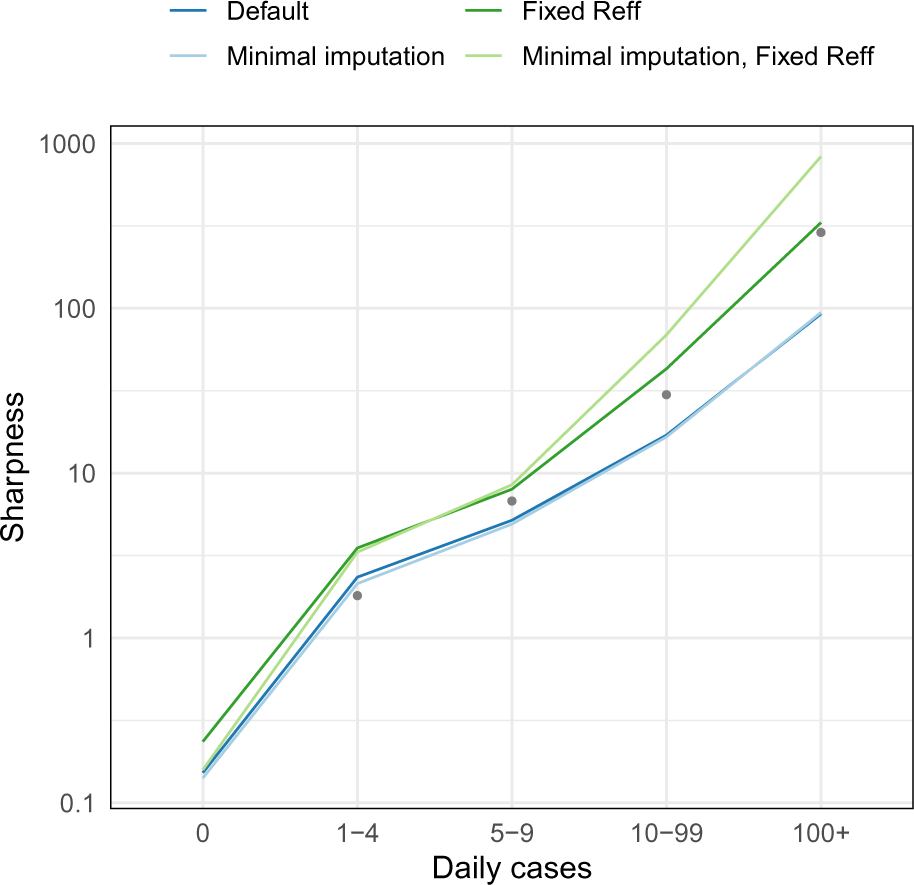
Forecast sharpness binned by the number of cases ultimately recorded for the day of the prediction. Mean recorded case numbers for each (non-zero) bin are shown as grey points.

**Figure S2:**
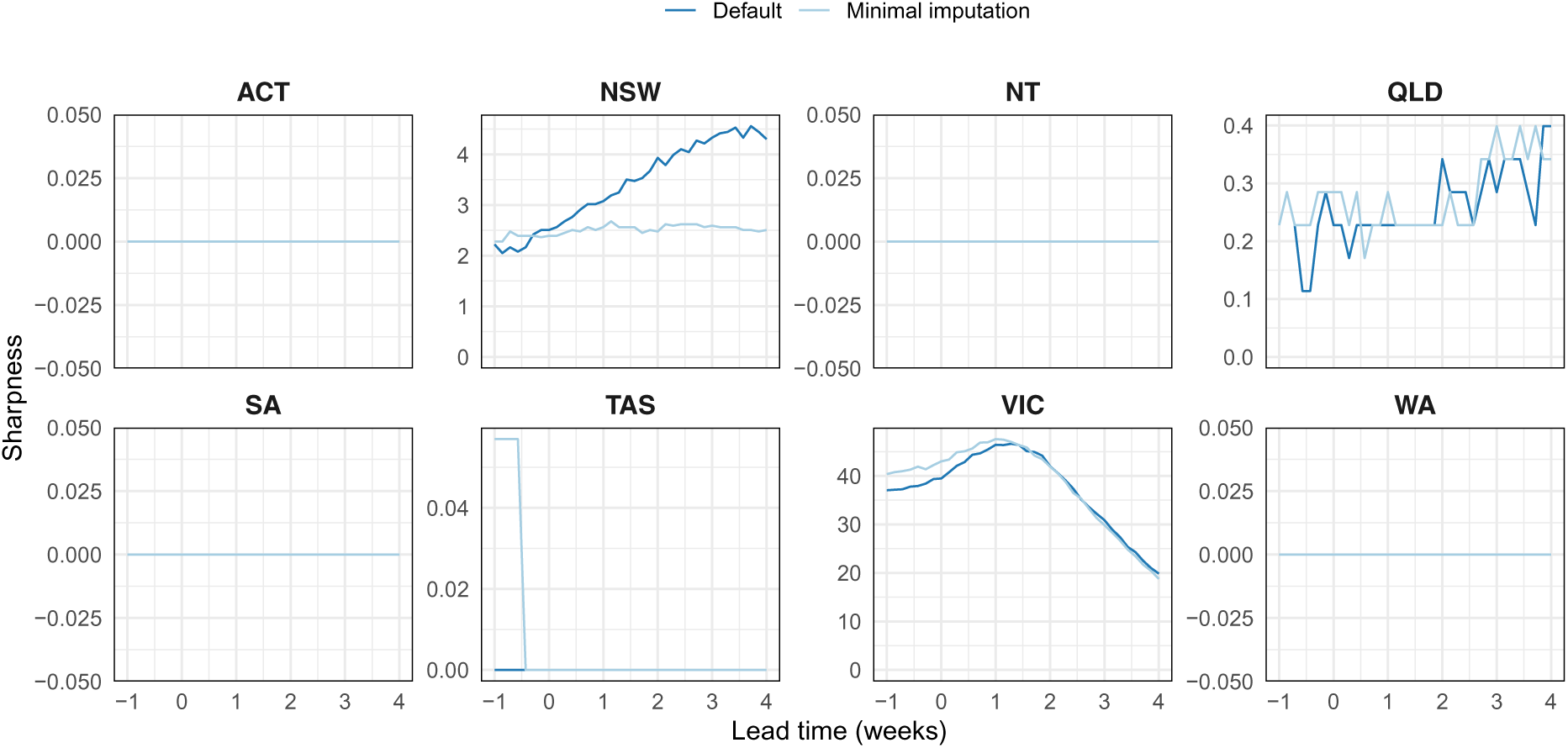
Forecast sharpness by lead time for the default and minimal imputation models, where the *R*_eff_ distribution gradually trends back towards the transmission potential (TP). During the second wave in Victoria, strong public health measures and social restrictions markedly reduced the TP, which caused forecasts to become sharper at longer lead times.

As a general rule, forecasts usually grow more uncertain as they predict further ahead [50]. However, the forecasts presented here for longer lead times tended to exhibit little growth in uncertainty, or even grow more certain (Figure S2).

**Figure S3:**
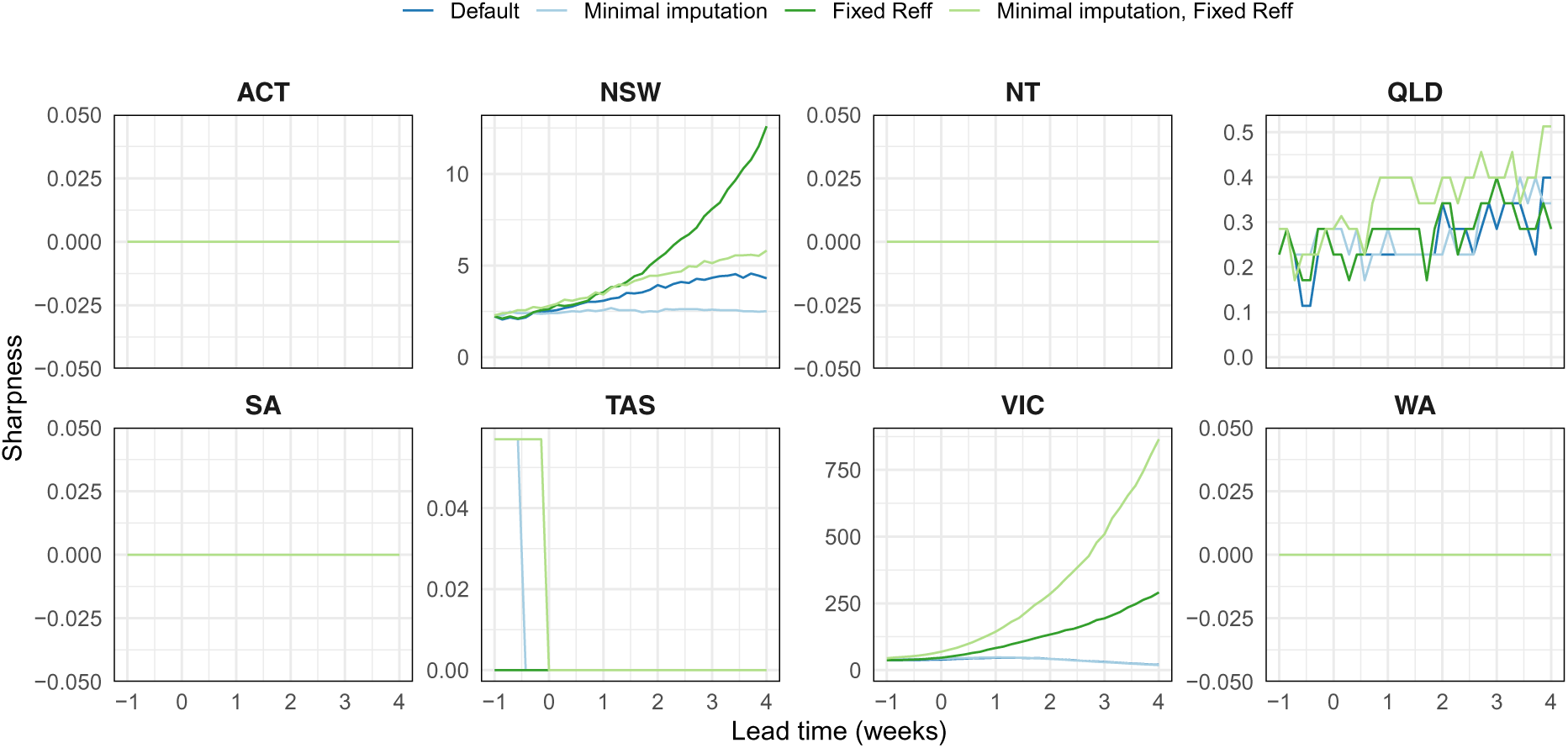
Forecast sharpness by lead time, shown separately for each forecast model.

For jurisdictions that primarily reported zero cases on a given day, the absence of epidemic growth meant that the forecast trajectories tended to local extinction and did not allow for a subsequent rebound. Accordingly, forecast sharpness in these jurisdictions remained very close to zero (i.e., no uncertainty) for all lead times.

For jurisdictions that reported substantial COVID-19 activity, forecasts for longer lead times either (a) exhibited little increase in uncertainty (New South Wales, Queensland); or (b) grew more certain but did not approach zero (Victoria). These jurisdictions imposed intense mobility and gathering restrictions in response to local COVID-19 activity, and the reduced contact rates in the general population meant that the model reproduction number tended to decrease below unity over the forecast horizon.

The forecasts did, however, grow more uncertain as the daily number of cases increased (Figure S1). And forecasts generated under the assumption that transmission would be sustained at the current active-case *R*_eff_ grew more uncertain for longer lead times (Figure S3, green lines).

### B.2 Forecast bias

**Figure S4:**
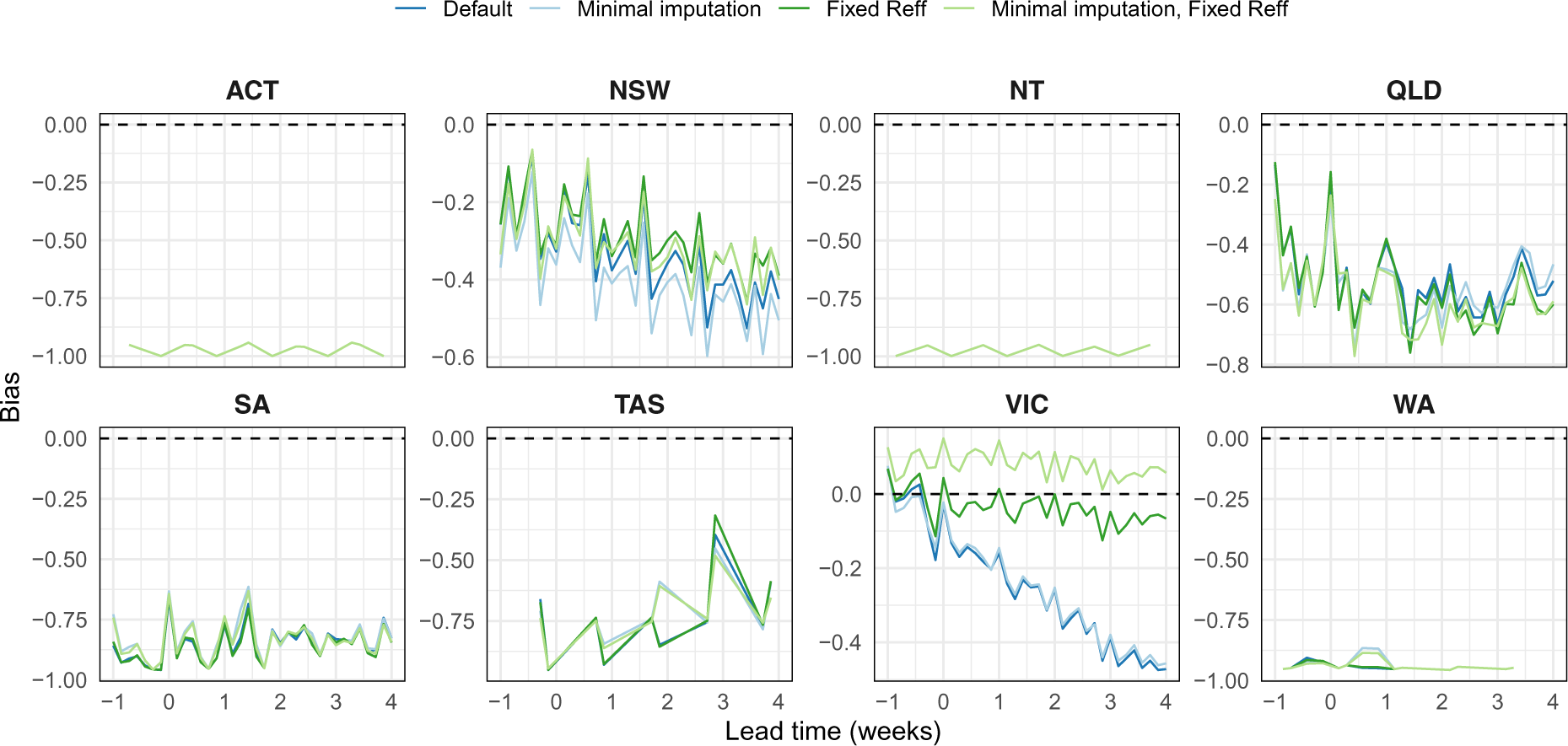
Forecast bias by lead time, for days where at least one case was reported.

**Figure S5:**
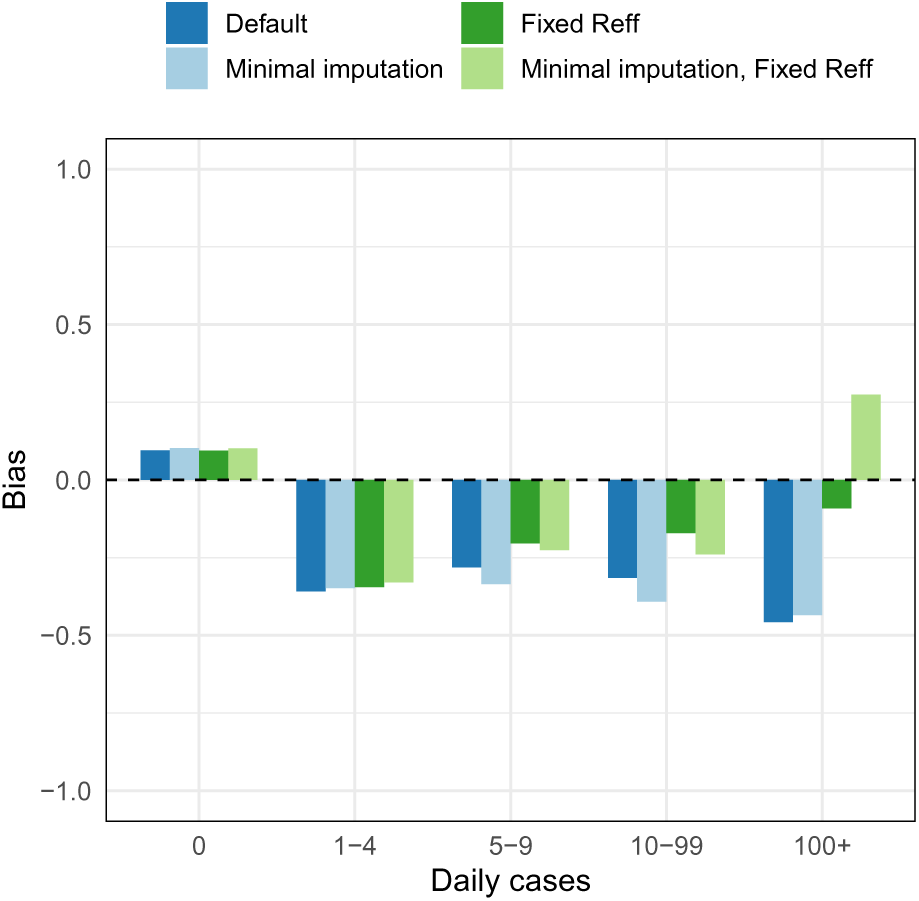
Forecast bias binned by the number of cases ultimately recorded for the day of the prediction.

Bias was defined so that 0 indicated an unbiased forecast, whose probability mass was distributed equally above and below the data, *−*1 indicated a forecast that completely under-estimated the data, and 1 indicated a forecast that completely over-estimated the data.

For jurisdictions that primarily reported zero cases on a given day (e.g., Australian Capital Territory, Northern Territory, South Australia, Tasmania, Western Australia) the forecast bias for days where one or more cases were reported is close to negative one (Figure S4). This indicates a deliberate model feature: our careful separation of the *potential* for reintroduction from the *consequence* of reintroduction (Section 4.3). For jurisdictions which had achieved local elimination and for which there was no evidence of reintroduction, the forecasts did not allow for subsequent epidemic activity.

**Figure S6:**
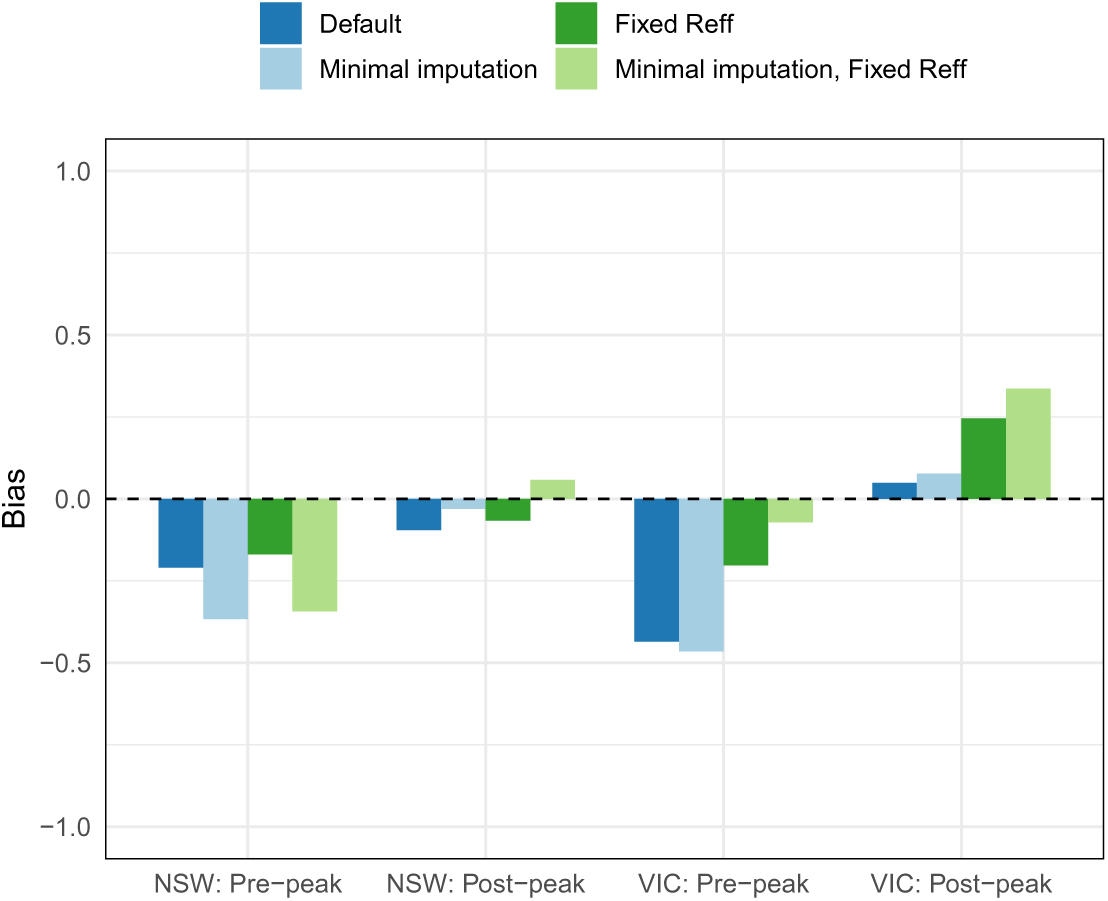
Forecast bias for the second wave in New South Wales and Victoria, reported seperately for data dates before and after the observed peak in each state (NSW: 29 July; VIC: 27 July).

In jurisdictions that saw substantial COVID-19 activity (New South Wales, Queensland, Victoria) the forecast bias was also negative and grew in magnitude as the lead time increased, but did not approach negative one (Figure S4). Forecasts exhibited a small positive bias on days where zero cases were reported (Figure S5, left-most category), because the forecasts always included some uncertainty and did not allow negative case counts. As the number of daily cases increased, the forecast bias became negative and grew in magnitude (Figure S5). When we consider the pre-peak and post-peak intervals of the second wave in Victoria and in New South Wales — the only period where any jurisdiction reported more than 100 local cases in a single day — we find that the forecast bias was negative prior to the peak and was approximately zero after the peak (Figure S6 and Figure S7). This suggests that the negative bias was due to a combination of factors.

**Figure S7:**
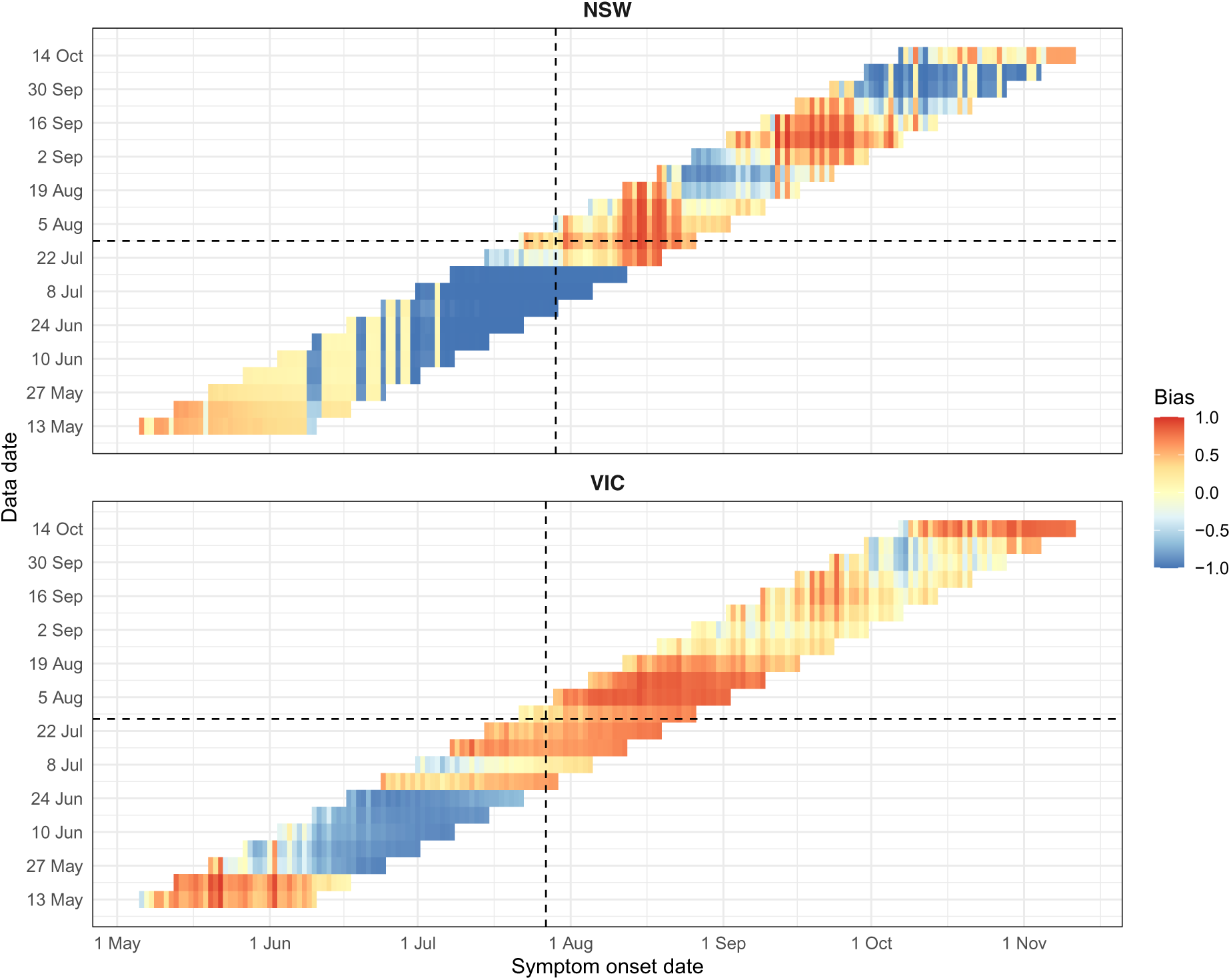
Forecast bias for the second wave in New South Wales and Victoria, shown separately for each day in the forecast horizon (x-axis) for each data date (y-axis). Dashed lines indicate the timing of the peak in each jurisdiction (NSW: 29 July; VIC: 27 July). There is a general trend to underestimate case counts (negative bias, blue) prior to the peak and to overestimate case counts (positive bias, red) after the peak. However, forecasts made in the few weeks prior to the observed peaks also exhibit positive bias.

During the second wave, the active-case *R*_eff_ was consistently higher than the whole-population transmission potential. So while the restrictions imposed during the second wave did reduce the active-case *R*_eff_ over time, and eventually brought the second wave to an end, our model over-estimated the decrease in local transmission over the forecast horizon. Despite this negative bias, the model consistently out-performed forecasts generated under the assumption that local transmission would be sustained at the current active-case *R*_eff_ (see Section B.3 for details).

### B.3 CRPS skill scores

#### B.3.1 Forecast skill by reported daily cases

**Figure S8:**
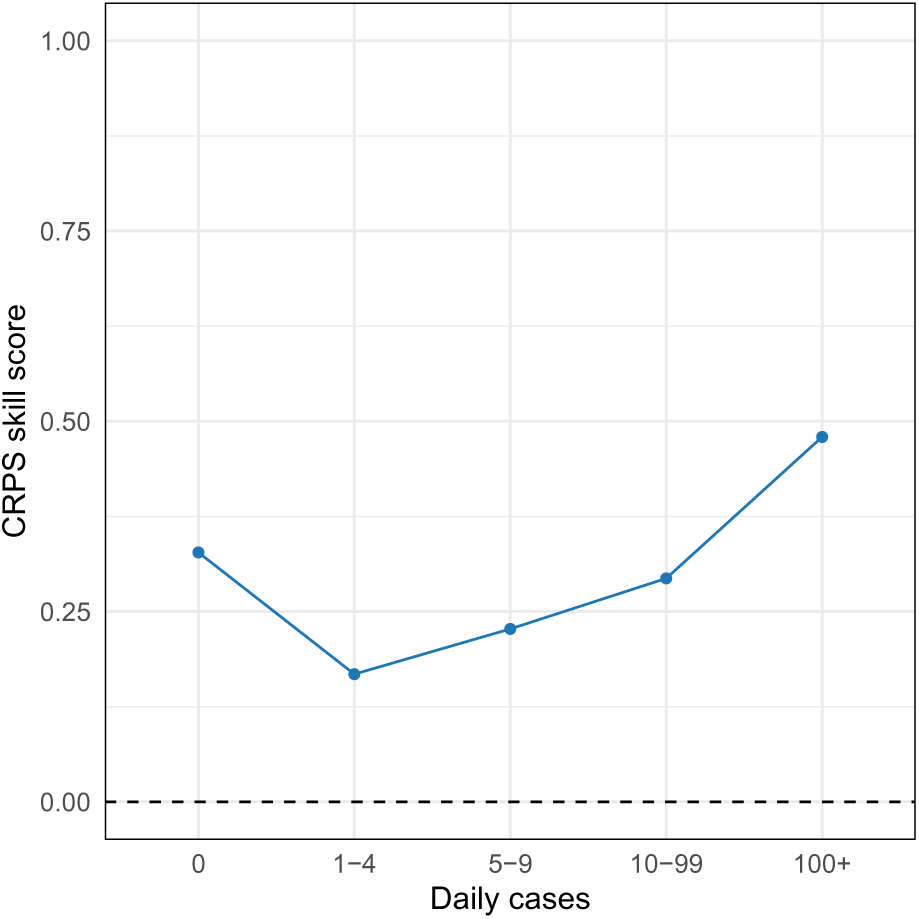
CRPS skill scores binned by the number of cases ultimately recorded for the day of the prediction.

We considered forecast skill as a function of the number of cases ultimately recorded on each day in each jurisdiction (comprising 1,688 observations). We divided the daily case numbers into five bins: 0 cases (*n* = 1298, 76.9%); 1–4 cases (*n* = 186, 11.0%); 5–9 cases (*n* = 57, 3.4%); 10–99 cases (*n* = 91, 5.4%); and 100+ cases (*n* = 56, 3.3%). The forecasts always out-performed the historical benchmark. The forecast skill across all jurisdictions was highest when daily case numbers were zero or ≥ 10, and was lower for days where 1–9 cases were reported (Figure S8). By using our model to predict how transmission would change over the forecast horizon (relative to the current active-case *R*_eff_ estimate) forecast performance was greatly increased (Figure S9, compare blue lines to green lines).

**Figure S9:**
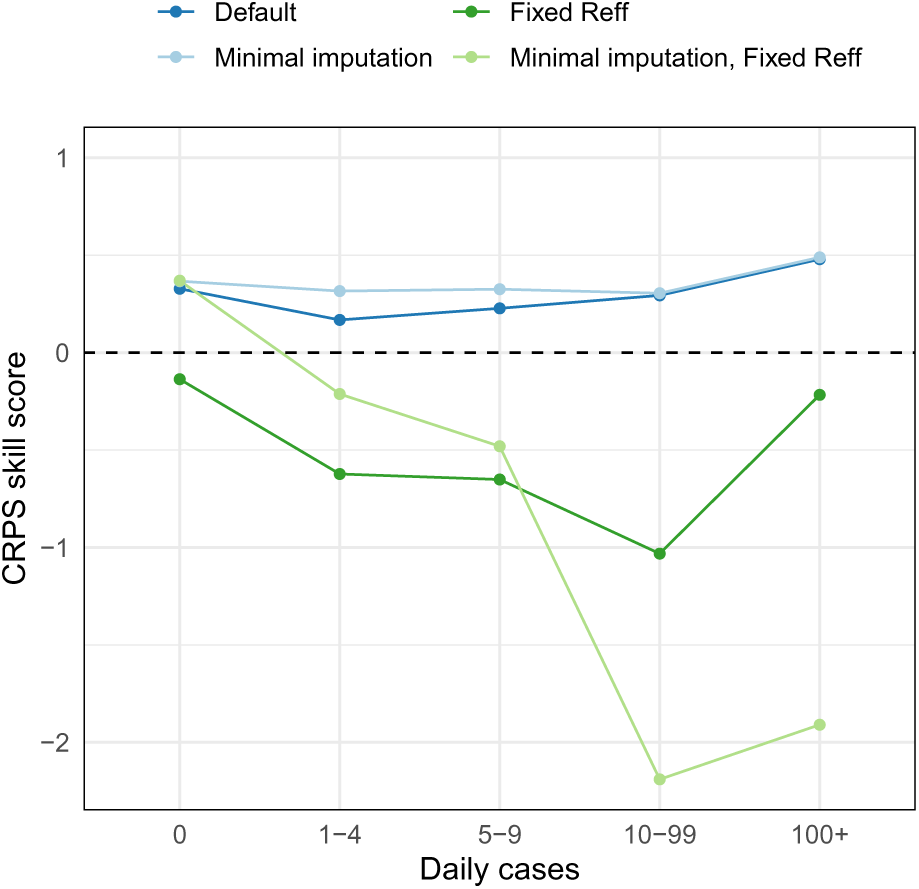
CRPS skill scores binned by the number of cases ultimately recorded for the day of the prediction, shown separately for each forecast model. The default model is slightly out-performed by the minimal imputation model when there are 1–9 cases per day, but has near-identical performance for higher case numbers and when there are zero cases.

#### B.3.2 Forecast skill when conditioning on incomplete data

**Figure S10:**
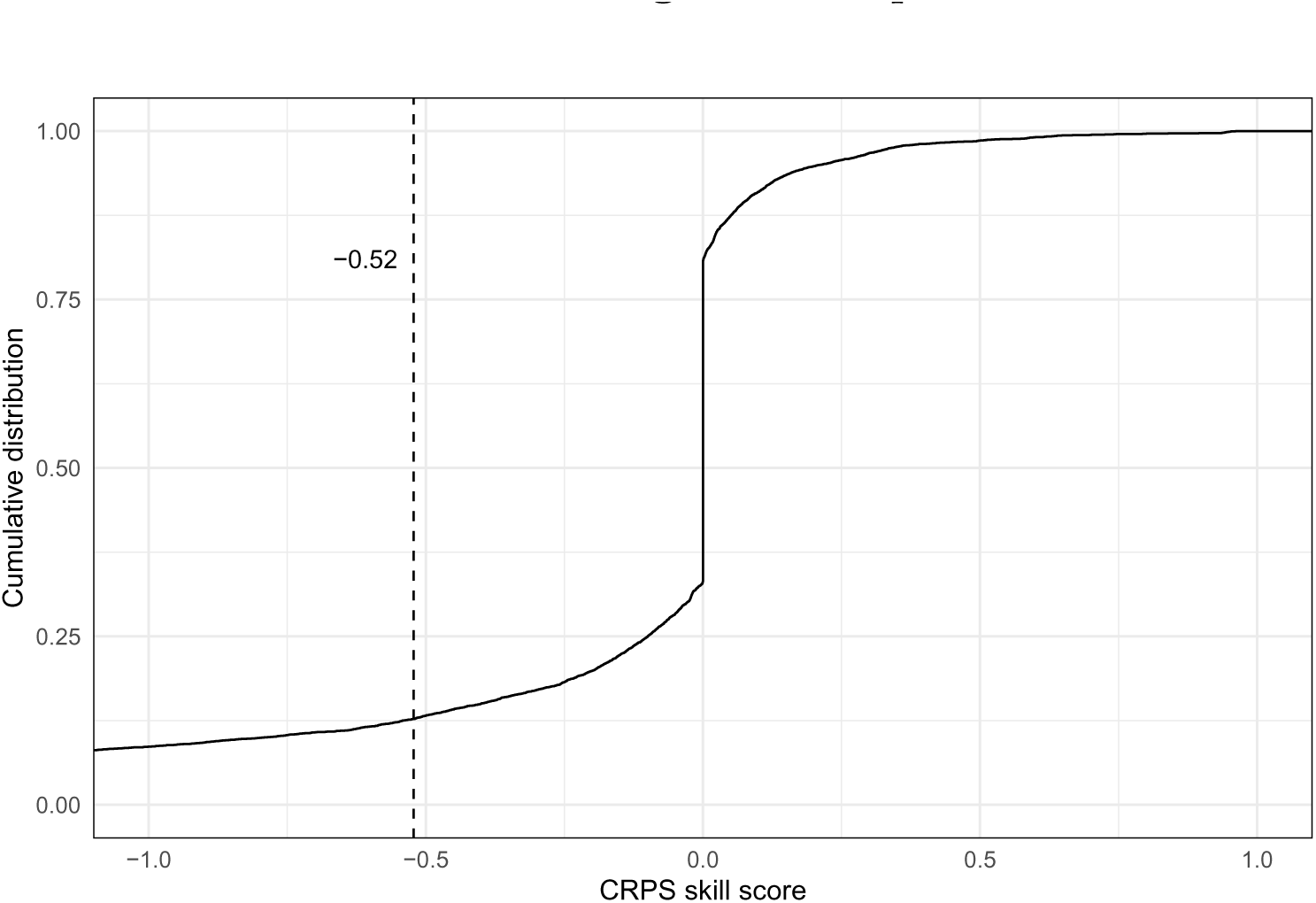
The cumulative distribution of CRPS skill scores for the minimal imputation model (which was not conditioned on data for dates where the detection probability was *<* 95%) relative to the default model. The mean skill score was -0.52 (dashed vertical line).

We estimated the delay distribution from symptom onset to case notifications being reported, and imputed the symptom onset data for cases where this field was not yet completed (see Section 4.1). The forecasting model was then conditioned on case counts, after accounting for under-reporting, for those dates where at least 50% of cases were estimated to have been reported (see Section 4.2). To determine how this treatment of the case data affected forecast skill, we used a separate “minimal imputation” model that was only conditioned on case counts for dates where at least 95% of cases were estimated to have been reported. We calculated CRPS skill scores for the “minimal imputation” model, relative to the original model, for each of the 7,488 predictions (8 jurisdictions, 26 data dates, 36-day forecasting window). The cumulative distribution of these skill scores is shown in Figure S10.

The minimal imputation model out-performed the default model for 19.3% of the predictions (median skill score of 0.09), yielded identical performance for 47.6% of the predictions, and was out-performed by the default model for 33.2% of the predictions (median skill score of -0.32). Identical performance primarily occurred on days with zero cases (98.1%), with the remainder occurring on days with 1–3 cases. The mean skill score was -0.52. This indicates that imputing symptom onset dates and accounting for delayed ascertainment provided additional information to that available in raw time-series data.

There were 25 predictions where the skill score was ≥ 90%, 23 of which pertained to a single data date for NSW (17 June) or Queensland (6 May). For New South Wales, the default model was conditioned on data for several days where 1 local case was reported, while the minimal imputation model was not conditioned on these data (because fewer than 95% of cases were estimated to have been reported for these dates). Accordingly, the default model predicted a subsequent increase in local cases, while the minimal imputation model predicted no local cases (95% CrI: 0–1 cases). These data were revised in subsequent data extracts, and each of the days with 1 reported local case now had no local cases.

The same explanation applies to Queensland, where only 2 cases were reported over the forecast horizon. The default model was conditioned on data for several days where 1 local case was reported, and predicted a larger increase in cases than the minimal imputation model, which was not conditioned on these data. Both sets of forecasts had identical medians (0 cases) and 50% CrIs (0–0 cases), but the default model had wider 95% CrIs (0–18 cases) than the minimal imputation model (0–4 cases).

#### B.3.3 Overall forecast skill in each jurisdiction

**Figure S11:**
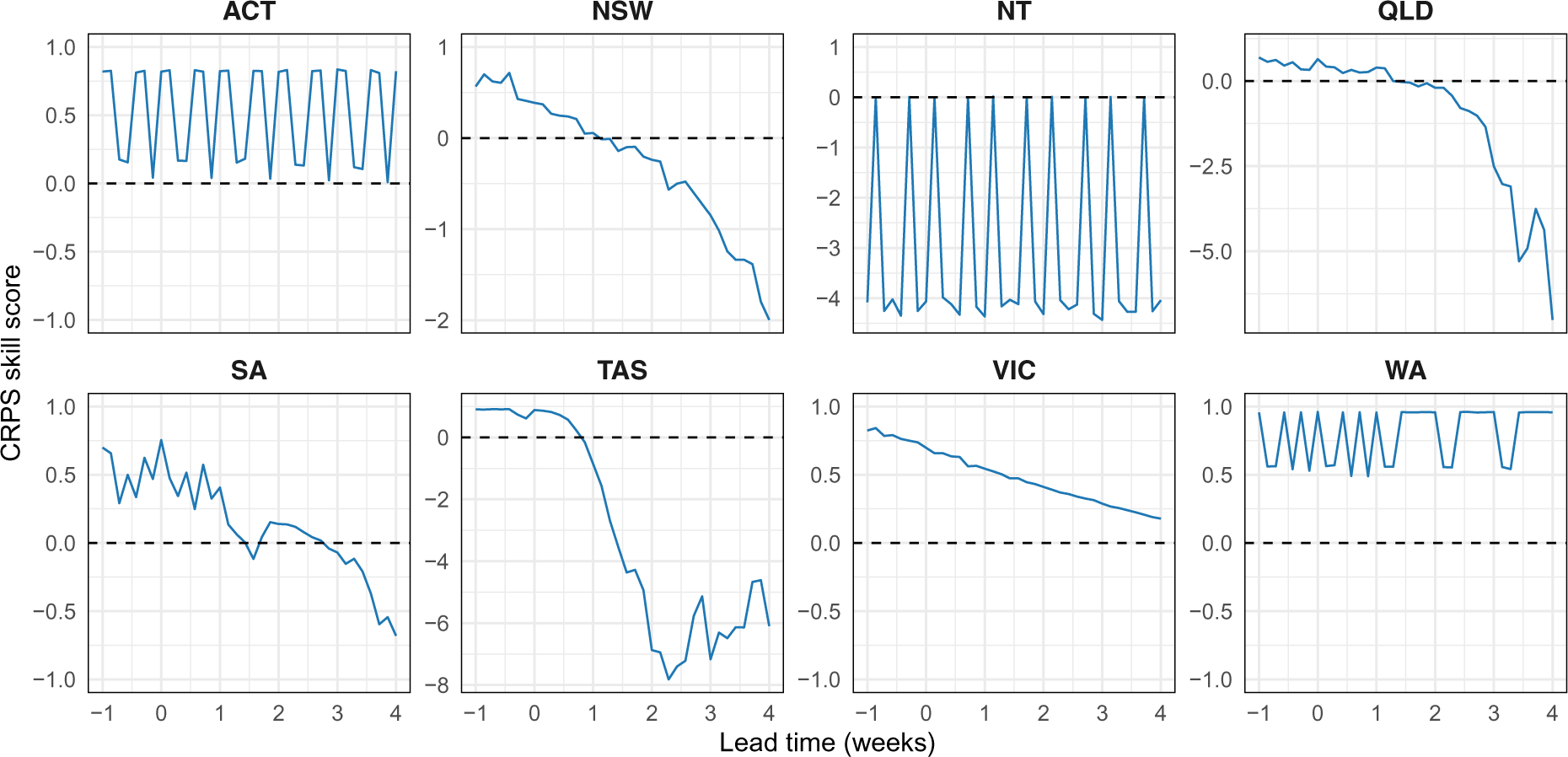
CRPS skill scores by forecast lead time, shown separately for each jurisdiction.

**Figure S12:**
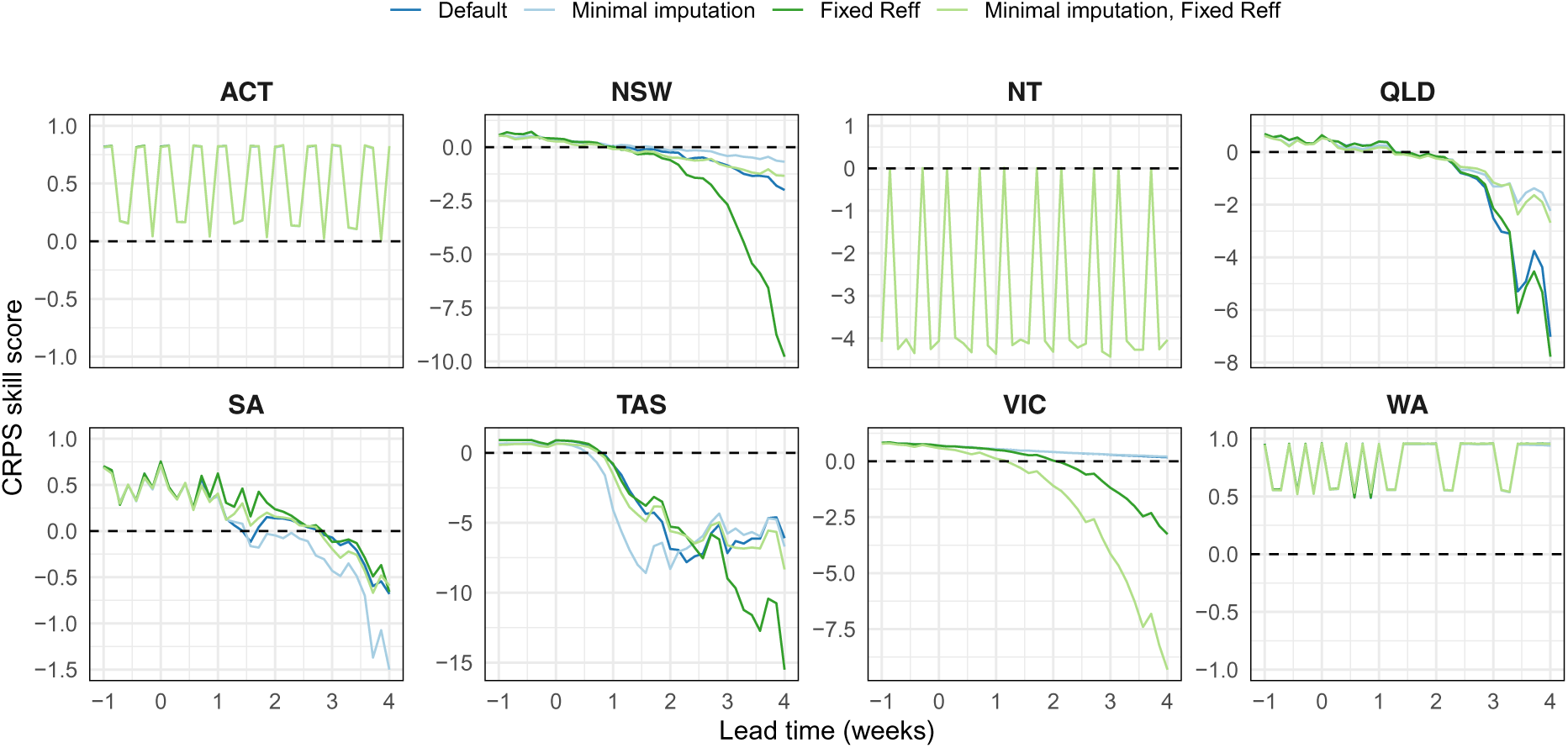
CRPS skill scores by forecast lead time, shown for each forecast model. Allowing the *R*_eff_ distribution to trend back towards the population mean (blue) consistently out-performed keeping the *R*_eff_ distribution fixed from the forecasting date onward (green). Conditioning on, and not conditioning on, the most recent case counts (dark shades and light shades, respectively) did not exhibit a consistent trend across jurisdictions.

We measured forecast skill relative to an historical benchmark forecast (see Section 4.4). Of the jurisdictions that primarily reported zero cases on a given day, the forecasts consistently out-performed the historical benchmark for all lead times in the Australian Capital Territory and Western Australia, and out-performed the historical benchmark for 1-week lead times in South Australia and Tasmania (Figure S11). The Northern Territory reported only three cases in the first wave and three cases in the study period (linked to separate importation events); these were ideal circumstances for the historical benchmark, which out-performed the model forecasts. For Tasmania, which experienced a local outbreak in the first wave and reported only two cases in the study period, the forecasts exhibited improved performance for lead times of up to one week, and exhibited decreased performance for longer lead times.

Of the three jurisdictions that experienced substantial COVID-19 activity (New South Wales, Queensland, Victoria), the forecasts exhibited improved performance for shorter lead times (Figure S11). The forecasts for Victoria, which experienced the largest local outbreak, substantially out-performed the historical benchmark at even the longest lead times. For New South Wales and Victoria, the model consistently out-performed forecasts generated under the assumption that local transmission would be sustained at the current active-case *R*_eff_ (Figure S12).

#### B.3.4 Forecast skill before and after the second wave peak

**Figure S13:**
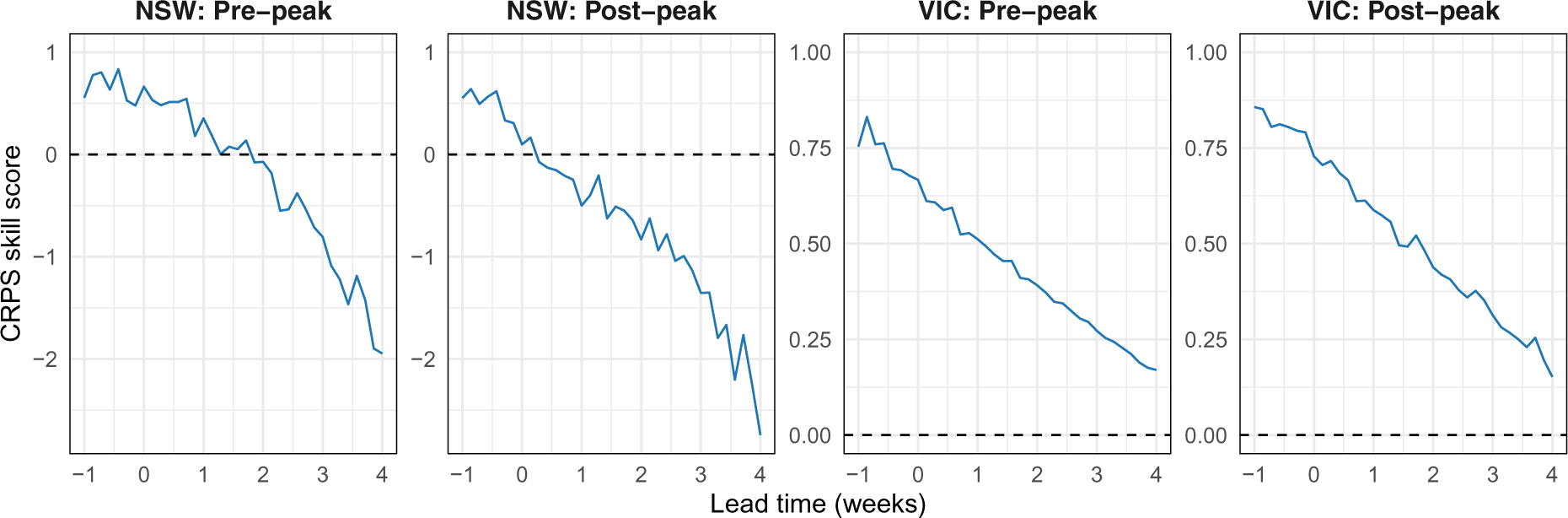
CRPS skill scores for the second wave in New South Wales and Victoria, shown seperately for data dates before and after the observed peak in each state.

**Figure S14:**
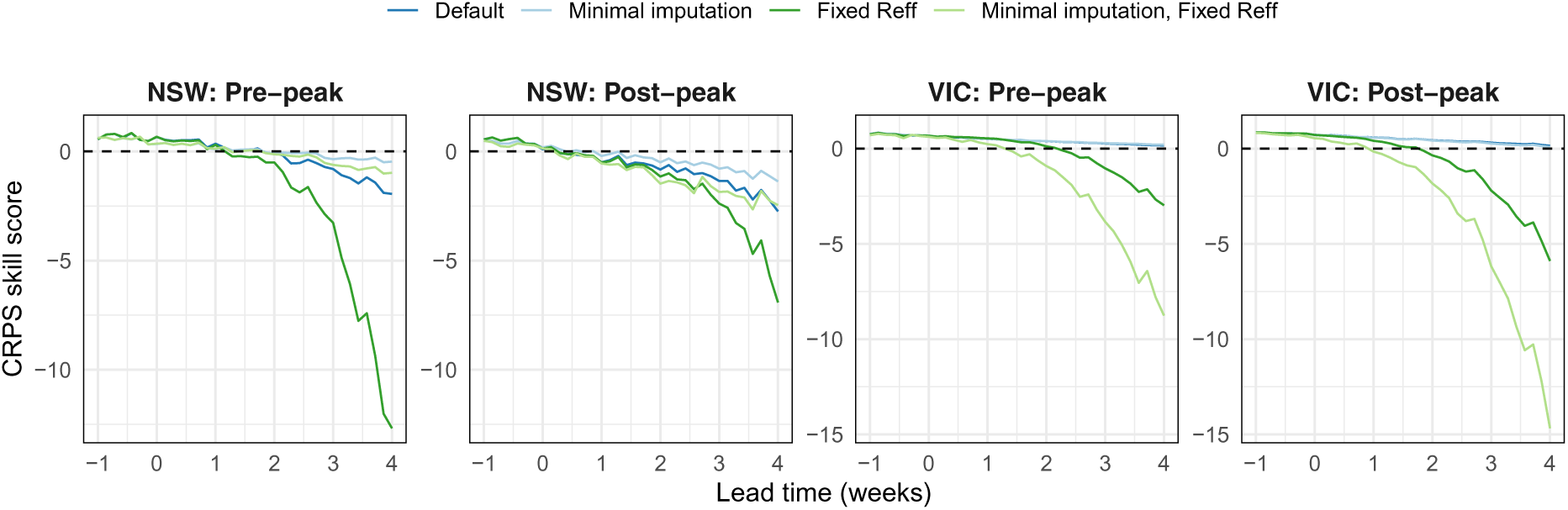
CRPS skill scores for the second wave in New South Wales and Victoria, shown seperately for each forecast model.

During the second wave (June to September, inclusive; 122 days) NSW reported 1–9 cases on 62 days (50.8%), and it was at these case levels that forecast skill was identified to be lowest across all jurisdictions (Figure S11). In contrast, Victoria reported 1–9 cases on only 14 days (11.5%), and reported at least 100 cases on 56 days (45.9%). Over this time period, the New South Wales forecasts out-performed the historical benchmark at shorter lead times (up to one week ahead), while in Victoria the forecasts consistently out-performed the historical benchmark at all lead times (Figure S13). By using our model to predict how transmission would change over the forecast horizon (relative to the current active-case *R*_eff_ estimate) forecast performance was greatly increased (Figure S14, compare blue lines to green lines).

#### B.3.5 Forecast skill by predicted local cases

**Figure S15:**
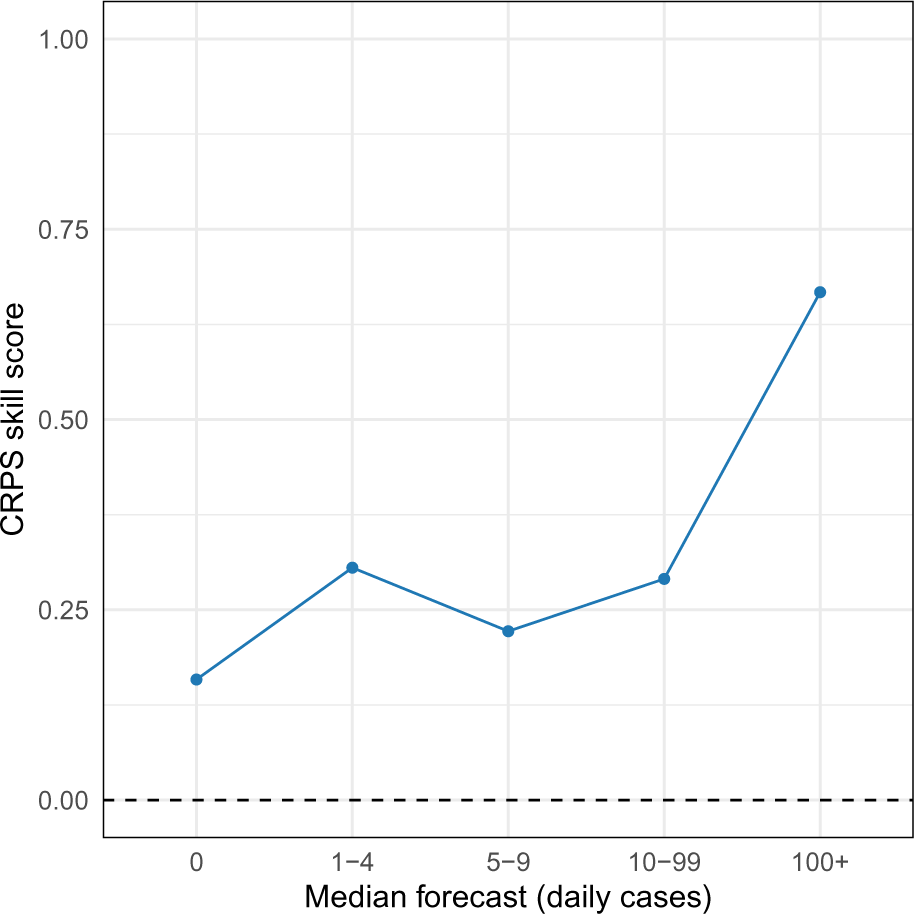
CRPS skill scores binned by the median forecast: 0 cases (81.2% of forecast days); 1–4 cases (6.9%); 5–9 cases (3.5%); 10–99 cases (5.5%); and 100+ cases (2.9%).

**Figure S16:**
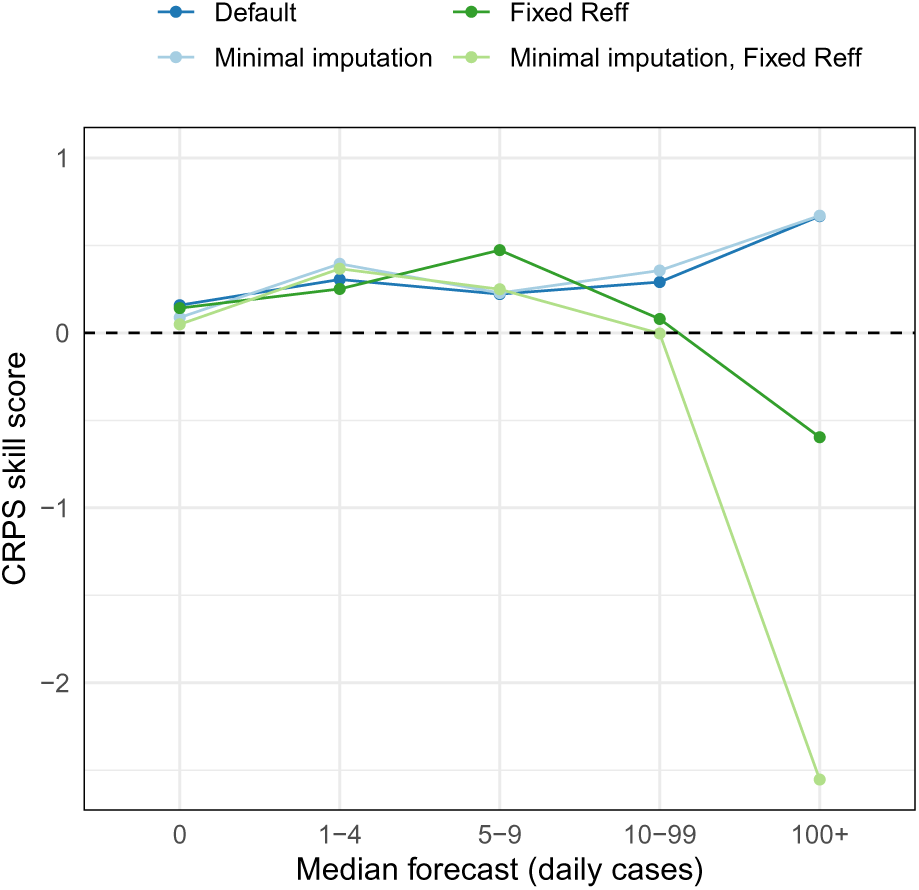
CRPS skill scores binned by the median forecast number of cases, shown separately for each forecast model.

When considering forecast skill as a function of the number of cases ultimately recorded for each day, as above, the results can only be evaluated in retrospect, since these numbers are unknown when the forecast is made. Accordingly, we also considered forecast skill as a function of the number of daily cases *predicted by the forecast model*, which may provide assistance when interpreting the forecast outputs in near-real-time. Our intent was to address the question “what does this forecast mean?” before the ground truth is known. The study period comprised 26 data dates across 8 jurisdictions, and the forecast interval spanned 36 days, resulting in 7,488 predictions of daily case counts. We binned these forecasts by the median predictions, which was correlated with forecast sharpness (Figure S1) and was therefore also indicative of forecast uncertainty, and used the same bins as before: 0 cases; 1–4 cases; 5–9 cases; 10–99 cases; and 100+ cases.

**Figure S17:**
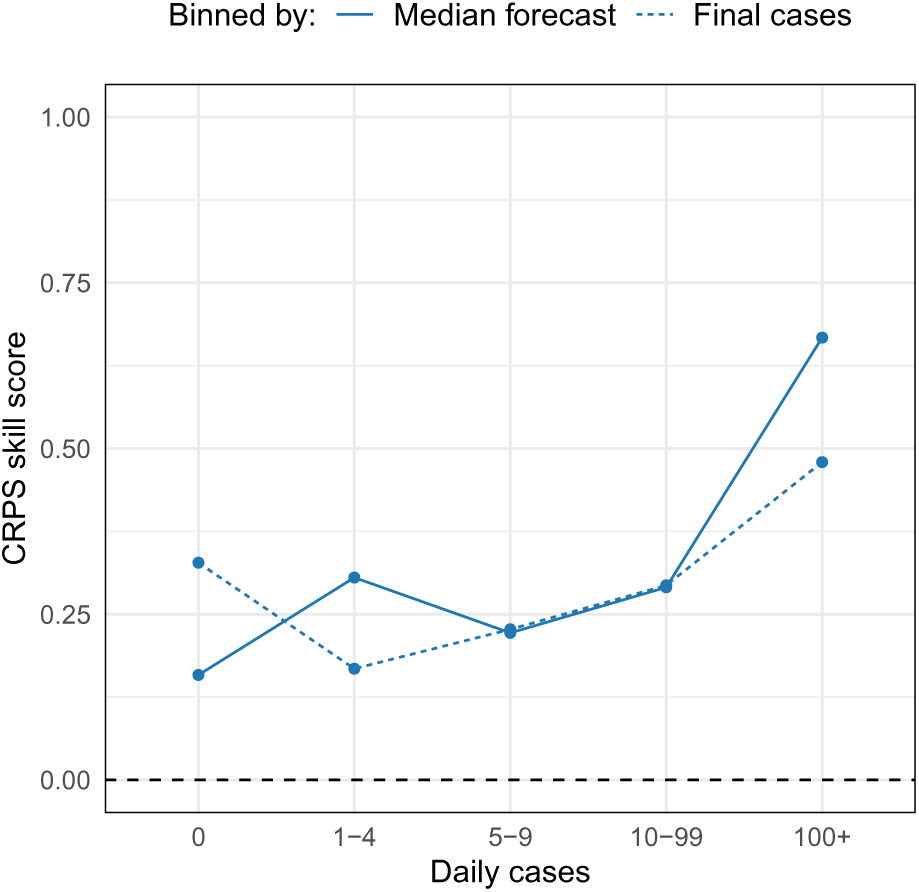
CRPS skill scores binned by the median forecast (solid line) and by the number of cases ultimately recorded (dashed line).

As per the skill scores aggregated by the number of cases ultimately recorded for each day, the forecasts always out-performed the historical benchmark. The forecast skill was lowest when the median prediction was zero cases, and was highest when the median prediction was *≥* 100 cases, followed by 1–4 cases (Figure S15). Forecasts generated under the assumption that transmission would be sustained at the current active-case *R*_eff_ exhibited similar skill to the default model forecasts when the predicted case numbers were low, but their skill decreased as the predicted case numbers increased (Figure S16).

In comparison to the skill scores aggregated by the number of cases ultimately recorded for each day, the skill scores aggregated by the predicted case numbers exhibit similar trends for *≥* 5 cases (Figure S17). In comparison to a median prediction of zero cases, the skill score is higher when there are zero reported cases. And in comparison to a median prediction of 1–4 cases, the skill score is lower when there are 1–4 reported cases. This suggests that when given a near-real-time forecast, where the future case counts are truly unknown, we can plausibly estimate the likely forecast skill when there is a moderate degree of local transmission.

### B.4 *R*_eff_ trajectories

**Figure S18:**
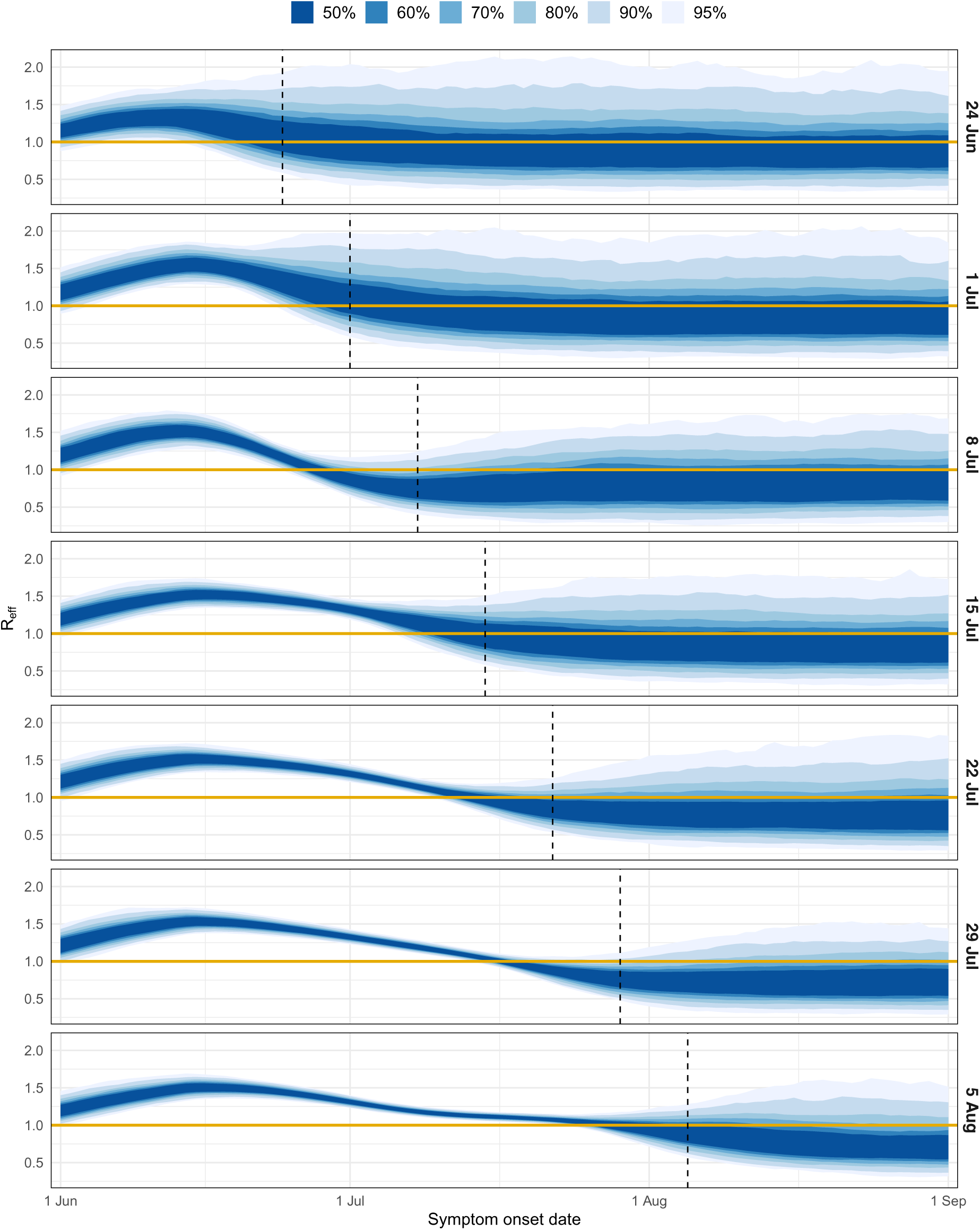
The *R*_eff_ trajectories for Victoria, shown separately for each data date (righthand axis labels). Each set of trajectories is shown as a spread of quantiles (blue shaded regions). Vertical dashed lines indicate the data date, and horizontal dark yellow lines indicate *R*_eff_ = 1.

**Figure S19:**
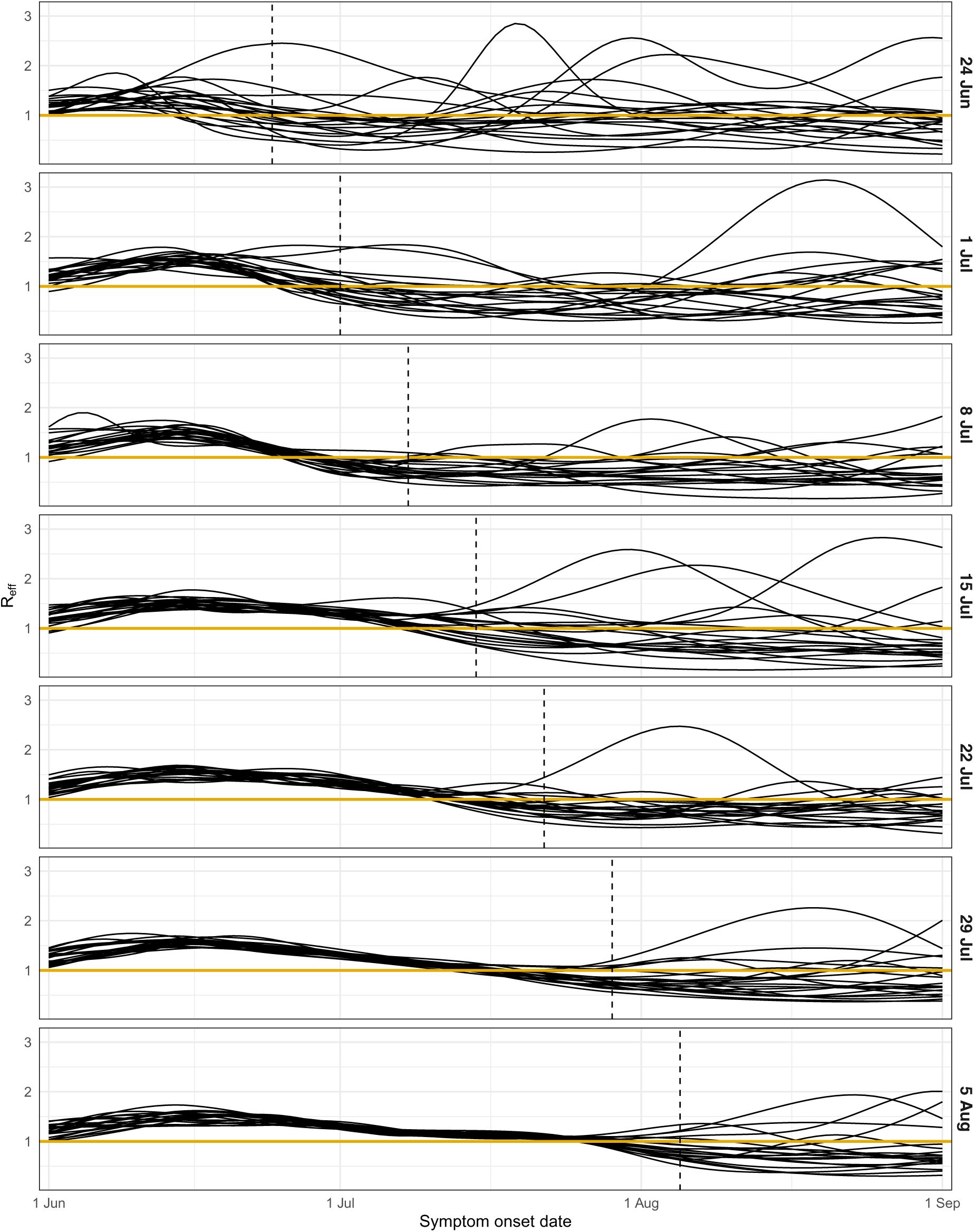
A sample of 20 *R*_eff_ trajectories for Victoria, shown separately for each data date (right-hand axis labels). Vertical dashed lines indicate the data date, and horizontal dark yellow lines indicate *R*_eff_ = 1.

### B.5 Healthcare-seeking behaviour and COVID-19 tests

**Figure S20:**
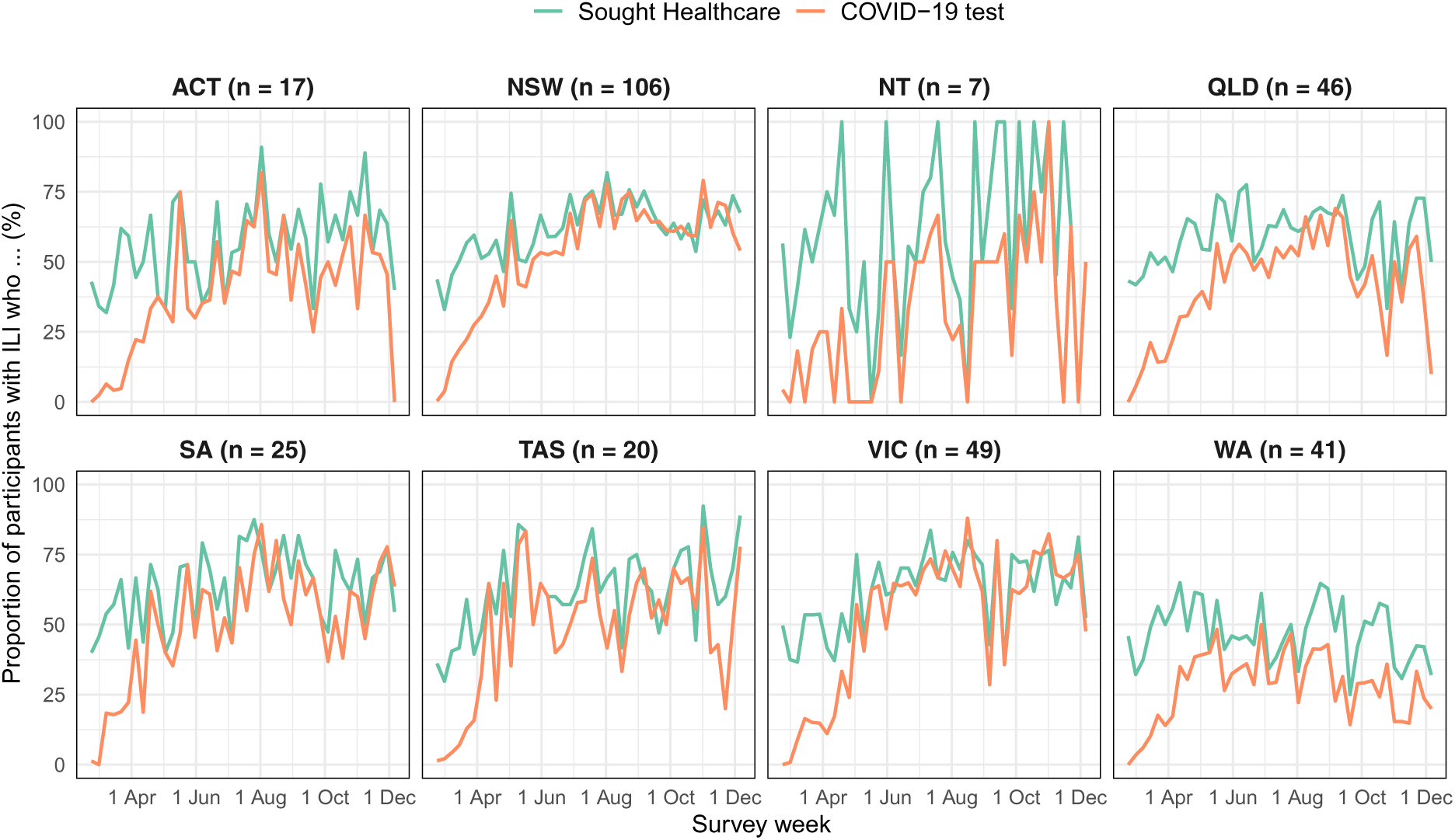
The proportion of Flutracking [32, 33] participants who reported ILI symptoms that (a) sought healthcare advice due to these symptoms (green lines); and (b) had a COVID-19 test (orange lines). The average number of participants *n* who reported ILI symptoms to Flutracking at each week is listed in the sub-plot captions. For comparison, in recent influenza seasons only 3–8% of participants who reported ILI symptoms had an influenza test [16].

